# A multilevel analysis of the prevalence and factors associated with multimorbidity in South Africa using 2016 Demographic and Health Survey data

**DOI:** 10.1101/2024.06.13.24308889

**Authors:** Matthew Hazell, Andre Pascal Kengne, Paramjit Gill, Dylan Taylor, Olalekan A. Uthman

## Abstract

Multimorbidity in Sub-Saharan Africa is under researched and includes distinct disease combinations to those seen in high income countries. The aim of this study was to determine the prevalence and distribution of multimorbidity in South Africa, as well as the associated individual, area-level and contextual factors. Multilevel logistic regression analyses were conducted on nationally representative 2016 South Africa Demographic Health Survey Data. The sample included 5,342 individuals (level 1) who completed the Adult Health questionnaire living in 691 neighbourhoods (level 2) from nine provinces (level 3). Multimorbidity was present in 44.6% of the study population and ranged from 36.8% in Gauteng to 52.8% in Eastern Cape. Individuals who were older, women, formerly married, black, obese, consumed a medium amount of sugary drinks, received education to primary or secondary school level or exposed to smoke at work had an increased risk of multimorbidity. Province level factors including poverty, rurality and unemployment, as well as neighbourhood level poverty were associated with multimorbidity. Some evidence of residual multimorbidity clustering was observed at the neighbourhood but not province level. Therefore, strategies that aim to tackle multimorbidity should address the risk factors identified and the wider determinants of health within neighbourhoods.

## Background

Individuals living with multimorbidity, defined as the co-existence of two or more chronic diseases, have a reduced quality of life, increased need to utilise healthcare, and greater mortality (Frølich et al., 2019; Loprinzi et al., 2016; Makovski et al., 2019; Nunes et al., 2016; Sum et al., 2019). Approximately 37% of the global population is believed to be multimorbid, with the burden primarily on the elderly (Chowdhury et al., 2023). Nevertheless, multimorbidity remains under researched compared to single conditions – this is particularly notable in low- and middle-income countries (LMICs) where only 5% of multimorbidity research originates (Xu et al., 2017).

Multimorbidity is increasing in LMICs, particularly in Sub-Saharan Africa (SSA), due to population ageing, lifestyle changes, and a changing climate and environment (Chowdhury et al., 2023). Additionally, multimorbidity in SSA is distinct from that of high-income countries due to the high prevalence of persistent infectious diseases (including human immunodeficiency virus (HIV) and tuberculosis), maternal and neonatal diseases, and injury related diseases co-occurring with non-communicable diseases (NCDs) (Oni et al., 2015). Concerningly, reports have also suggested that multimorbidity is appearing at younger ages in SSA (Oni et al., 2015). Therefore, research within this context is required to inform the specific healthcare management of multimorbidity in SSA.

South Africa is an upper-middle-income country within SSA, and one of its most urbanised and developed nations (McGranahan & Martine, 2012; World Bank, 2024). Many health inequalities exist in South Africa across socioeconomic, ethnic, and geographical divisions (Ataguba et al., 2011; Weimann et al., 2016). These inequalities are partly a legacy of the Apartheid era, which divided the population by race and deprived most South Africans of basic human rights, and the high levels of unemployment and urbanisation seen since the advent of democracy in 1994 (Chopra & Sanders, 2004). Indeed, poorer groups are reported to experience a higher burden of NCDs, and a higher prevalence of NCD risk factors including binge drinking and obesity, despite NCDs being viewed as diseases of affluence (World Obesity Federation, 2020).

There is considerable heterogeneity in prevalence estimates of multimorbidity in South Africa. A recent systematic review found estimates ranging from 3-23% in studies with younger people, and 30-88% in older adults (Roomaney et al., 2021). This is due to contrasting study designs and differing definitions of multimorbidity: definitions that capture more chronic conditions will result in a higher multimorbidity prevalence. Of the single conditions that make up multimorbidity, hypertension, anaemia, and HIV are believed to be the most prevalent (Roomaney et al., 2022).

Identified individual-level risk factors for multimorbidity in South Africa include sex, age, area of residence, occupation, education, income, marital status and body mass index (BMI) (Alaba & Chola, 2013; Garin et al., 2016; Roomaney et al., 2021). However, the literature is sparse and contradictory. The role of an individual’s area of residence on multimorbidity, which has been demonstrated in Ghana, is likely also important in South Africa and intra-context correlation needs to be considered (Kuuire et al., 2023). Higher burdens of multimorbidity have been reported in parts of KwaZulu-Natal and Eastern Cape and lower burdens in the provinces of Limpopo and Mpumalanga (Weimann et al., 2016). However, there has been no research into the extent of variation of multimorbidity by area in South Africa which is important to understand the influence of specific contexts on multimorbidity.

This study aims to determine the prevalence and distribution of multimorbidity in South Africa, as well as the individual, neighbourhood, provincial and contextual factors associated with multimorbidity. This research will inform policy makers to allocate resources appropriately, to develop prevention programmes and manage multimorbidity.

## Materials and methods

### Study design and data sources

A multilevel logistic regression analysis of South Africa Demographic and Health Survey (DHS) (SADHS) 2016 data was conducted. SADHS 2016 is a nationally representative cross-sectional household survey that provides information on demographic and health indicators.

The DHS programme has assisted with over 350 nationally representative household surveys across 90 countries since 1984 (Corsi et al., 2012; United States Agency for International Development, 2023). Surveys are usually conducted every five years per country, with topics tailored to relevant population health issues. The DHS is an important source of data for policy making, monitoring and evaluation in many LMICs.

The SADHS 2016 followed a stratified two-stage sample design (National Department of Health et al., 2019). Seven hundred and fifty primary sampling units (PSUs) were selected from the 26 sampling strata, based on urban, traditional and rural areas for each of the nine provinces in South Africa (there was no strata for traditional areas in Western Cape). A fixed number of 20 dwelling units (DUs) were randomly selected from each of the PSUs. This design permits estimates of key variables for the country, for each of the nine provinces, and of urban, rural and traditional areas. Data collection took place over six months, from 27 June 2016 to 4 November 2016.

All DUs were eligible for the primary modules on women, fertility and children, and half were subsampled for modules on men and adult health. The adult health module included self-reported chronic conditions and biomarker collection, for anthropometry, anaemia, hypertension, HBA1c levels for diabetes, and HIV, for those aged over 15.

All participants of SADHS 2016 completed consent forms. The anonymised datasets with necessary permissions were obtained from the DHS programme (https://www.dhsprogram.com/data/) for this secondary analysis, and no further ethical clearance was required.

### Study population

Men and women aged 18 and over were eligible for inclusion into this study if they contributed data to SADHS 2016 adult health modules. Individuals under 18 were excluded to ensure comparability to the literature. Individuals were also excluded from the study if they were missing information on the multimorbidity outcome.

### Variables

#### Outcome variable

Multimorbidity is measured by counting the number of co-existing chronic conditions, with a cut-off of at two or more conditions (Johnston et al., 2019). Twelve current chronic diseases contributed to the binary multimorbidity outcome (multimorbidity or no multimorbidity): tuberculosis, hypertension, stroke, high blood cholesterol, anaemia, chronic bronchitis, diabetes, asthma, cancer, heart disease, HIV, and chronic pain. These were recorded through self- and biomarker-reports. Unlike previous research, individuals missing any information on a biomarker reported disease were coded as missing rather than not having the disease, as this was believed to reduce misclassification bias (Roomaney et al., 2022).

In the adult health module, individuals were asked about the presence of several diseases (tuberculosis, stroke, hypertension, high blood cholesterol, chronic bronchitis, diabetes, asthma, cancer, heart disease, and chronic pain) and whether they took medication for the diseases. For example, participants were asked ‘Has a doctor, nurse or health worker told you that you have or have had any of the following conditions: heart disease?’, with participants responding either ‘Yes’, ‘No’ or ‘Don’t know’ (National Department of Health et al., 2019). A response of ‘Don’t know’ was recoded as ‘No’. Individuals who self-reported a disease did not always concurrently report taking medication for the disease. To ensure diseases were chronic, tuberculosis had to have occurred within the previous 12 months and chronic pain had to last longer than three months; these were confirmed with responses to further questions.

Biomarker-measured diseases (hypertension, anaemia, diabetes, and HIV) were collected from blood specimens from finger pricks and measured using validated instruments in consenting individuals.

Digital blood pressure monitors were used to take three blood pressure measurements, at intervals of three minutes or more (National Department of Health et al., 2019). As per convention, the study excluded the first blood pressure measurement and utilised an average of the remaining two measurements (Roomaney et al., 2022). Hypertension was classified as individuals who have either a systolic blood pressure of ≥140 mm Hg or a diastolic blood pressure of ≥90 mm Hg (World Health Organisation, 2023). People with non-hypertensive biomarkers who self-reported that they were on medication to treat hypertension were recorded as hypertensive. Implausible biological marker values were recoded as missing (SBP <69 or >271 mm Hg, DBP <29 or >151 mm Hg).

For anaemia, non-pregnant women, pregnant women, and men with haemoglobin levels below 7 g/dl, 9g/dl and 9 g/dl were classified as anaemic, respectively (National Department of Health et al., 2019). Anaemia testing was carried out on site and results were adjusted for smoking status and altitude. Dried blood spots analysed at the Global Clinical and Viral Laboratory in Durban were used to record diabetes and HIV. Individuals with HbA1c ≥ 6.5 mmol recorded by a blood chemistry analyser were classified as diabetic, as were those with healthy HbA1c values who self-reported taking diabetes medication. An enzyme-linked immunosorbent assay (ELISA) was used to test for HIV, a second ELISA and an alternative confirmatory rapid test (Bio-Rad) were used to confirm the initial ELISA test. Individuals were either classified as HIV positive, HIV negative or inconclusive, those who were inconclusive were recoded as missing.

#### Explanatory variables

Included explanatory variables were informed by literature review and a-priori reasoning, all were self-reported or derived from self-reported variables. The socioeconomic variables included were household wealth index, education level, occupational status, health insurance and marital status. Individual level health variables included BMI (missing for women who were pregnant or had been in the last two months), dietary health, sugary drink intake, smoking status, alcohol drinking and exposure to smoke at work. Two variables representing access to old and new media, respectively, were included as a proxy for access to health information. Finally, variables for age, sex and ethnicity were also included. Those who answered ‘don’t know’ were recorded as missing information on that variable. Further information on how these variables were derived and coded can be found in Table S1 in the Supplementary file.

Neighbourhoods were defined as respondents from clusters of households which serve as the PSU within the DHS. Poverty, rurality, and unemployment level were chosen as the neighbourhood and province level explanatory variables, with illiteracy only investigated at the neighbourhood level. These were defined as the proportion of individuals living in the most deprived group for each neighbourhood and province (proportion in the lowest wealth category, living rural or traditional, unemployed and illiterate). This was split into three categories (low, medium and high), with neighbourhood rurality as a binary variable, and calculated with the larger adult-health sample (N= 9,512) to include more contextual information.

### Statistical methods

To describe and compare the characteristics of individuals in the study population with and without multimorbidity, descriptive statistics were produced which summarised the included covariates in those with and without multimorbidity. A description of the data by province was also produced. To identify the conditions contributing most to the burden of multimorbidity the prevalence of each chronic condition in the study population was recorded. All analyses were conducted using STATA 18 and descriptive analyses were adjusted for sample weight, stratification and clustering as per DHS recommendations (StataCorp, 2023).

To analyse the individual/ household (level 1), neighbourhood (level 2) and province (level 3) level factors associated with multimorbidity, multivariable multilevel logistic regression models were produced. Multilevel analysis has been utilised in much prior social epidemiology research when investigating hierarchical data and considers the individual probability of the outcome to be statistically dependent on area of residence (Benebo et al., 2018; Bolarinwa et al., 2022; Consolazio et al., 2021; Due et al., 2009; Ijaiya et al., 2022; Liyew & Teshale, 2020). The method is ideal to discover the determinants of multimorbidity, their differences across hierarchical levels and the magnitude of the clustering effects at these levels, for an accessible introduction see (Merlo, Chaix, et al., 2005a, 2005b; Merlo et al., 2006; Merlo, Yang, et al., 2005). Five multilevel models were produced. Model 1 contained no covariates to demonstrate the variance in the outcome variable attributed to clustering at the neighbourhood and province level. Model 2, 3 and 4 included individual-level, neighbourhood-level and province-level variables only respectively. Finally, model 5 was fully adjusted and included all individual-level, neighbourhood-level and province-level variables. This method allows an investigation into how much of the neighbourhood and province level differences in the outcome were explained by individual, neighbourhood and province level characteristics.

Adjusted odds ratios with 95% confidence intervals were reported for fixed effects associations between individual, neighbourhood and province level variables and the outcome. Measures of area-level variance from random effects, were the median odds ratio (MOR) and variance partition coefficient (VPC). The MOR estimates the variance in multimorbidity expressed as an odds ratio attributed to neighbourhood and province contexts and represents the extent to which the individual probability of multimorbidity is determined by residing in a neighbourhood and province. The MOR can be conceptualised as the median increased risk of multimorbidity from moving to an area with a higher risk (Larsen & Merlo, 2005; Merlo et al., 2006). The VPC represents the proportion of response variance at the neighbourhood and province levels of the model. A Chi-squared test was used to determine the strength of evidence for variation at the neighbourhood and province level.

The number of observations with missing data for each variable was reported. A complete case analysis was conducted which is valid if missingness is, conditional on the covariates, independent of the outcome which seemed likely (e.g. consent to measure BMI may be less likely to be given for obese patients) (White & Carlin, 2010).

The effects of excluding individuals who contributed to the adult health module of SADHS 2016 but were missing information on chronic diseases that contributed to the outcome was investigated. Missingness of each chronic disease, demographic information of those missing outcome data and missingness by province were summarised. Additionally, a sensitivity analysis was conducted using this larger sample, including individuals missing information on chronic diseases, to account for missing outcome data with multiple imputation using chained equations (Rubin, 2004). The data was imputed 25 times based on the missing at random (MAR) approach, which assumes that the expansive information available on demographics and other diseases predict most of the variation in missing data (Little & Rubin, 2019). Data checks were made following imputation to ensure pregnant or recently pregnant women’s BMI was missing. The statistical analysis plan was repeated with this imputed sample (N=9,512 weighted).

## Results

### Characteristics of the study population

*Figure 1* shows the study population selection process (N=5,650, weighted sample N=5,342). Of the 15,292 households selected for the SADHS sample, 11,083 (83%) were successfully interviewed. 12,717 individuals were identified for the adult health subsample, and 10,336 completed interviews (81%) of whom 9,468 (91.6%) were aged 18 or over and 5,650 (59.6%) with complete outcome data were included in this analysis.

**Figure 1.**
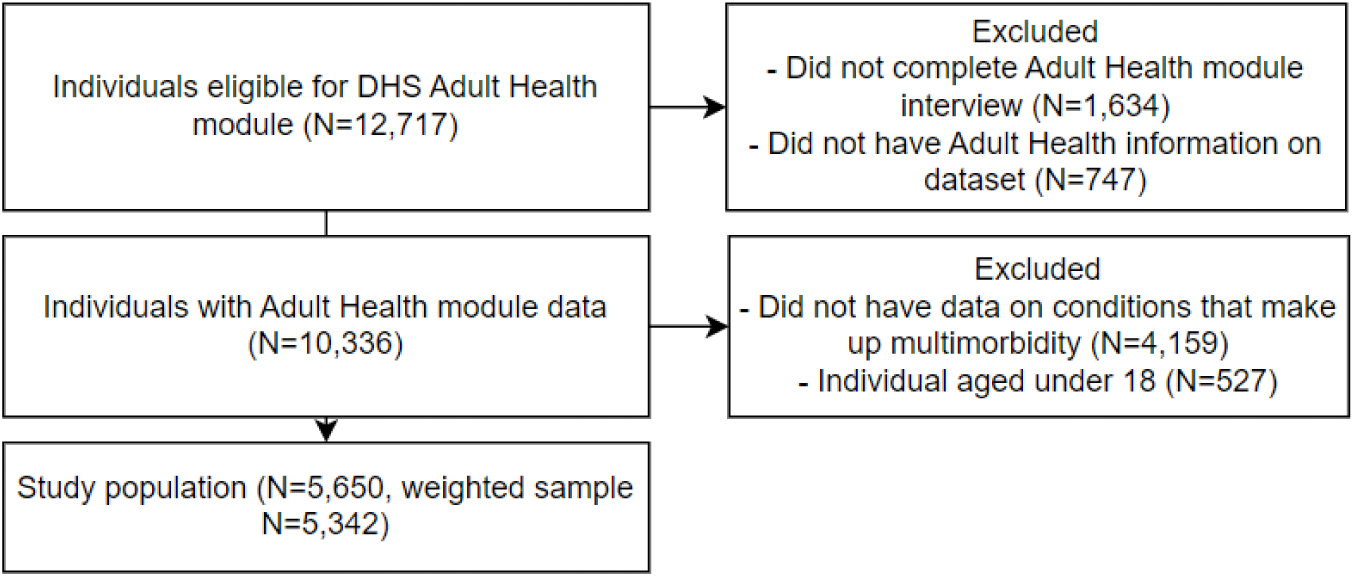
Study population selection process.

### Descriptive data

The study population adjusted for sample weight, stratification and clustering included 5,342 adults (level 1), living in 691 neighbourhoods (level 2) from nine provinces (level 3) in South Africa. The baseline characteristics of the study population are described in Table 1. Multimorbidity was present in 44.6% (N=2,382) of the study population, with women having a higher prevalence than men (51.4% and 33.1%, respectively). The mean age of adults in this sample was 41.6 (SD 17.6) years, individuals with multimorbidity were substantially older. The majority of the participants were black (86.4%), never smokers (75.8%), unemployed (64.9%), and received education up to secondary level (62.6%). Individuals with and without multimorbidity were balanced in terms of wealth and ethnicity. Healthy diets, lower sugary drink intake, never smoking and never drinking were more common amongst those with multimorbidity, but multimorbid individuals were more likely to be obese.

**Table 1.**
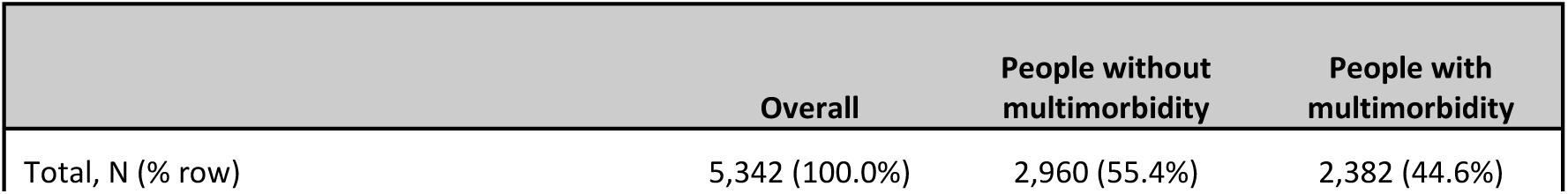
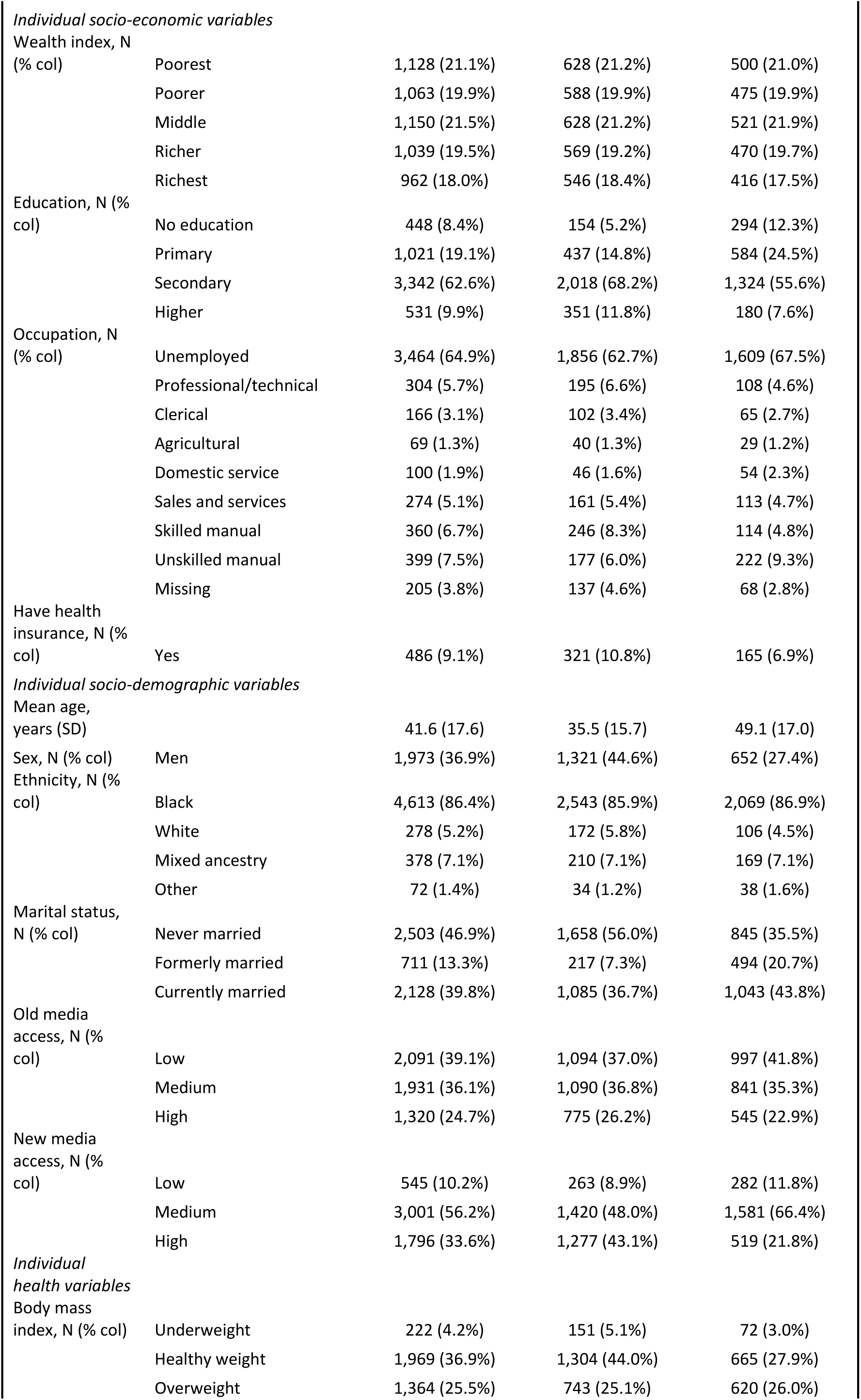
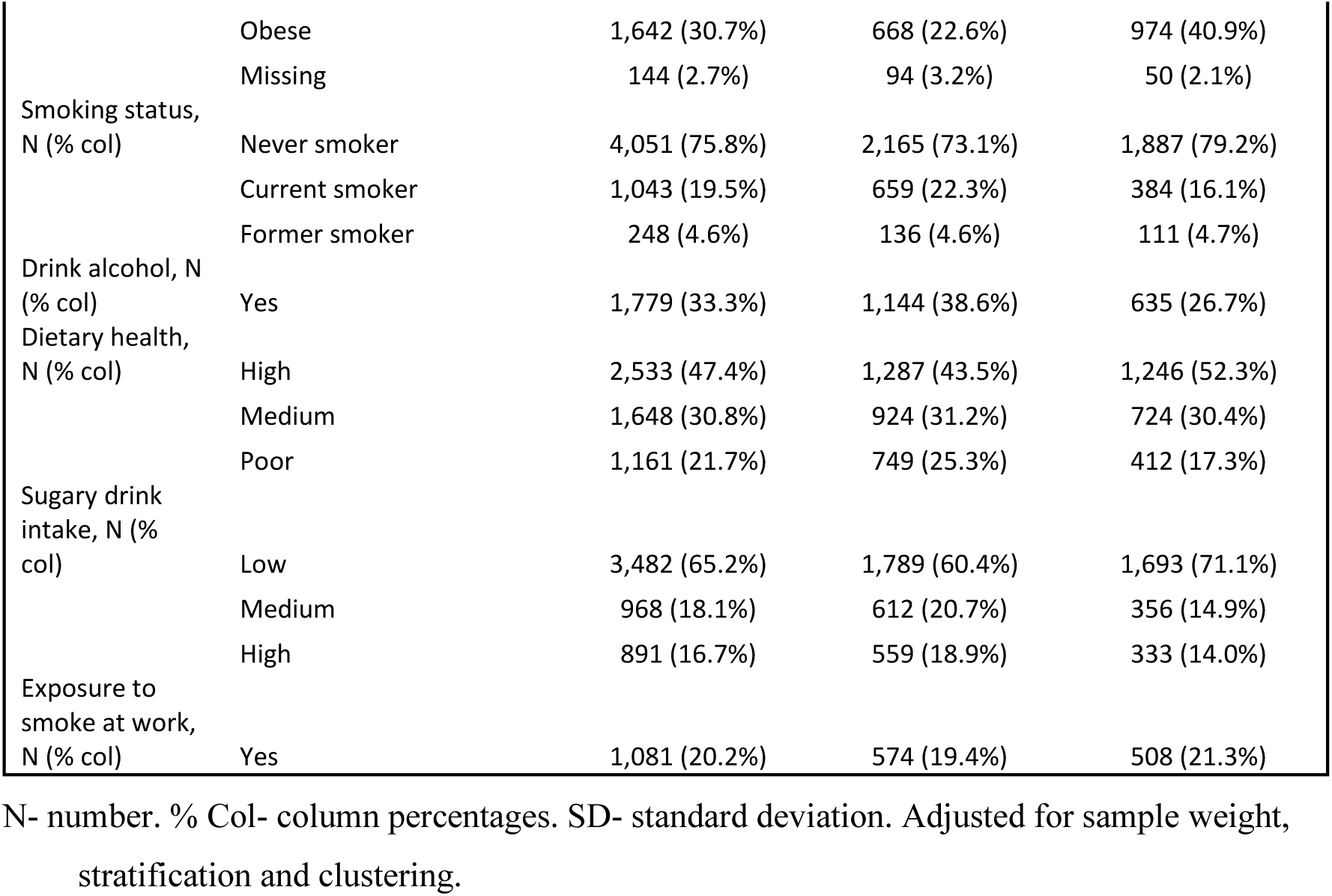
Characteristics of the study population by multimorbidity status.

Table 2 presents a description of the data by province. Multimorbidity was highest in the province of Mpumalanga (52.2%) and lowest in Limpopo (37.0%). There was large variation in province level factors. Western cape contained the oldest individuals and had the lowest proportion of people living in the poorest wealth group (4.7%), a rural area (3.7%), unemployment (55.4%) and illiteracy (4.9%). By contrast, 38.9% of those living in the Eastern Cape belonged to the poorest wealth group and 70.7% were unemployed whilst, 85.0% of those from Limpopo lived in rural areas and 17.9% were illiterate.

**Table 2.**
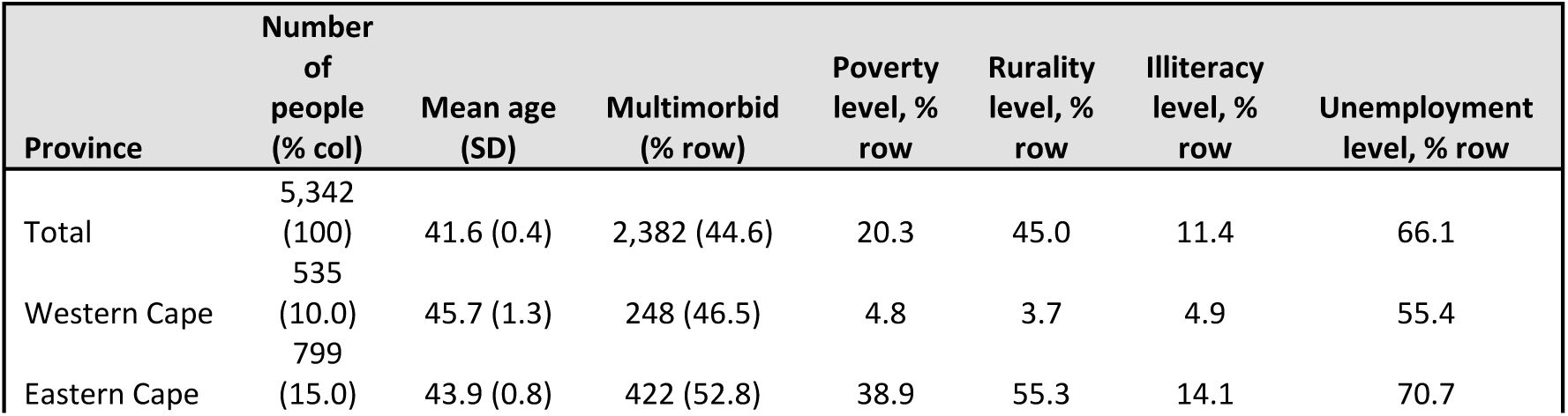
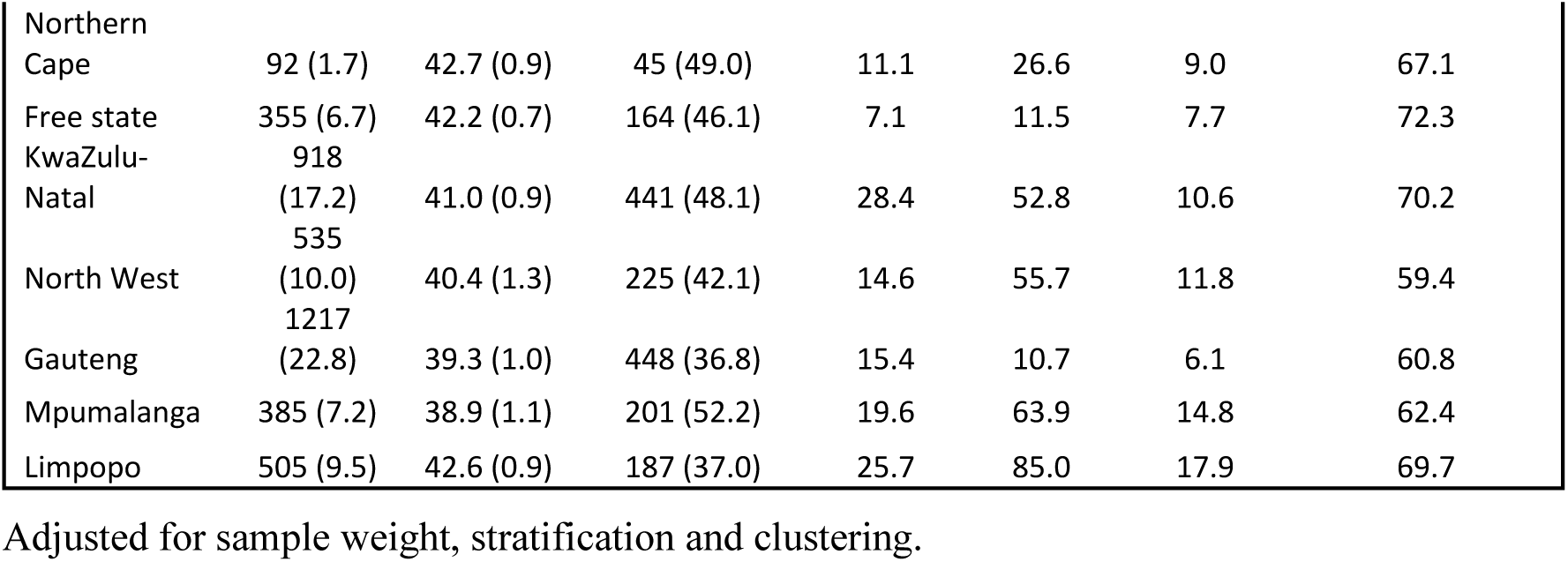
Characteristics of the study population by province.

Almost half of the study population (46.4%) were prevalent for hypertension (See Figure 2). Around one fifth had chronic pain, HIV, diabetes and anaemia, whilst few reported cancer (1.2%), chronic bronchitis (1.4%), stroke (1.4%), and tuberculosis (1.5%).

**Figure 2.**
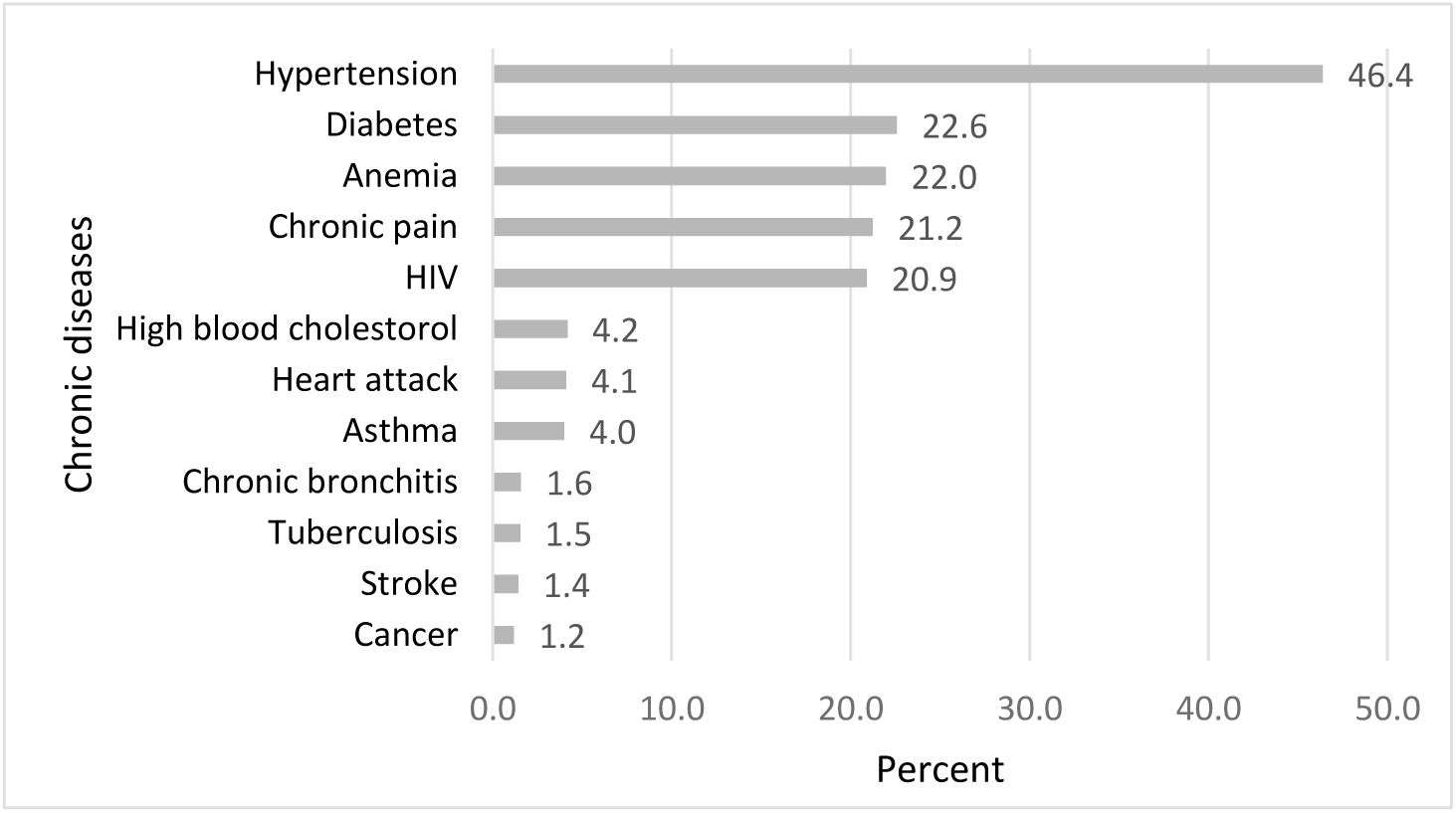
Prevalence of individual chronic diseases in the study population. Adjusted for sample weight, stratification and clustering.

### Fixed effects (measures of association)

Table 3 presents the results from the five multilevel logistic regression models constructed. Fixed effects results reported in text are from model 5, which is adjusted for all individual, neighbourhood and province level variables.

**Table 3.**
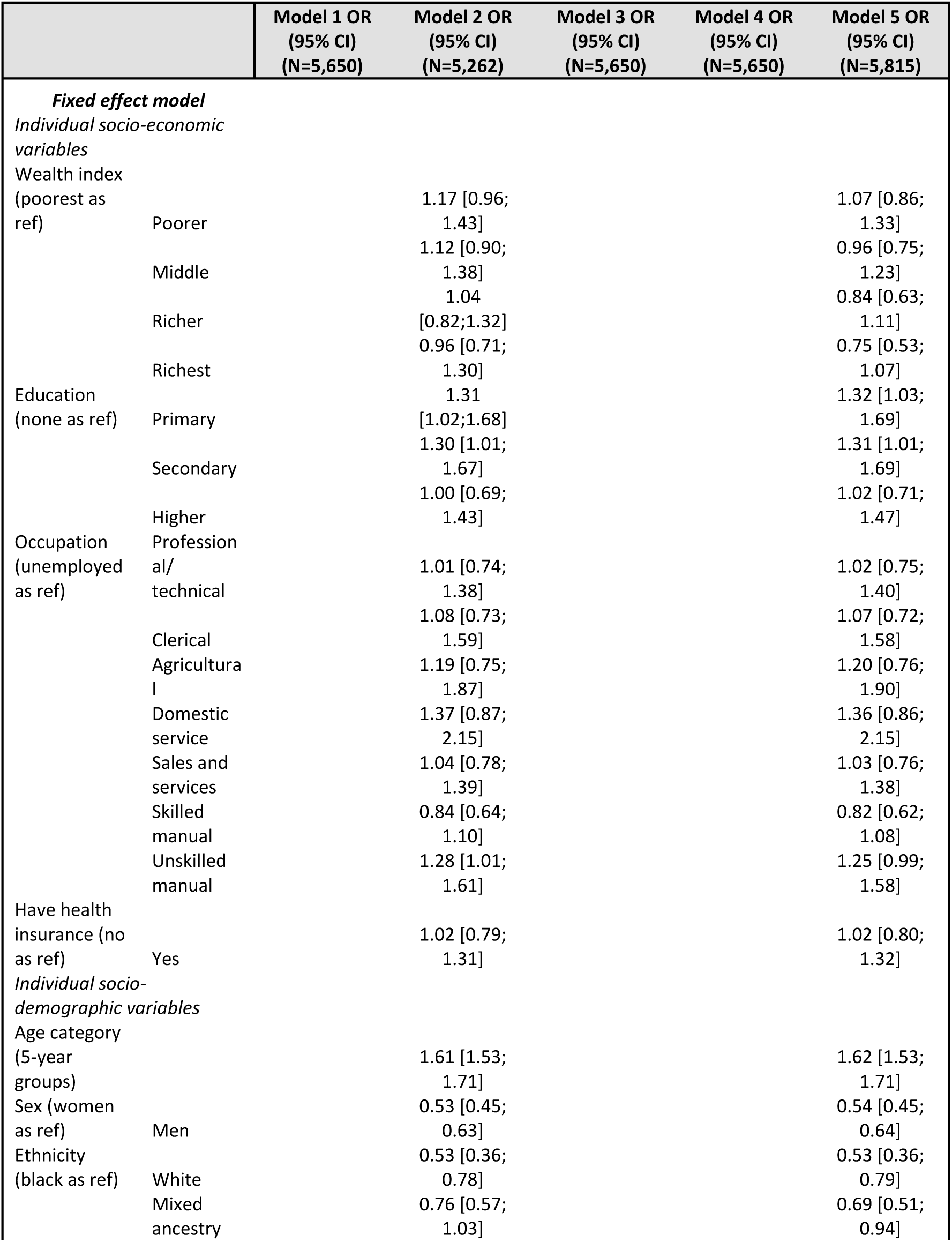
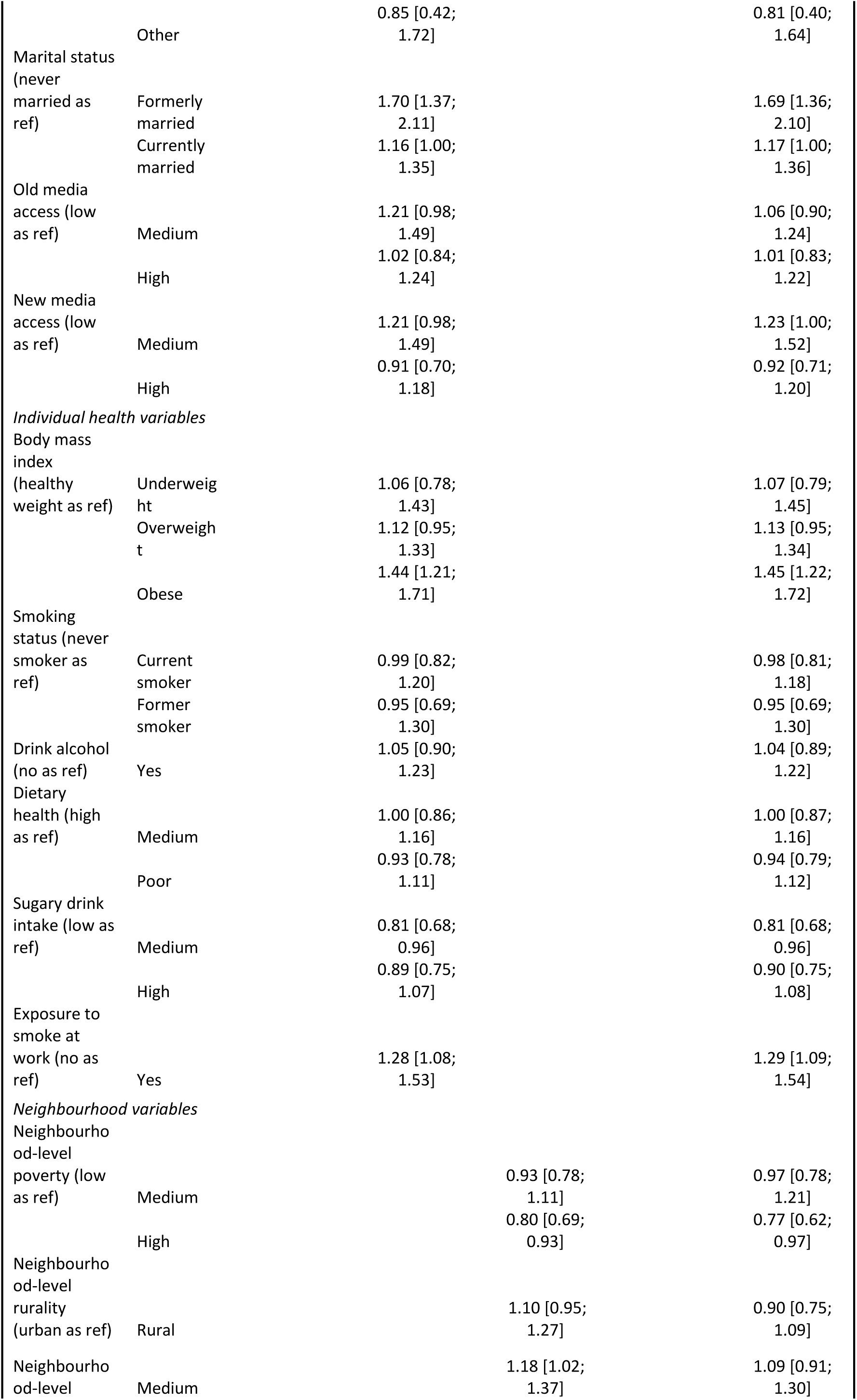
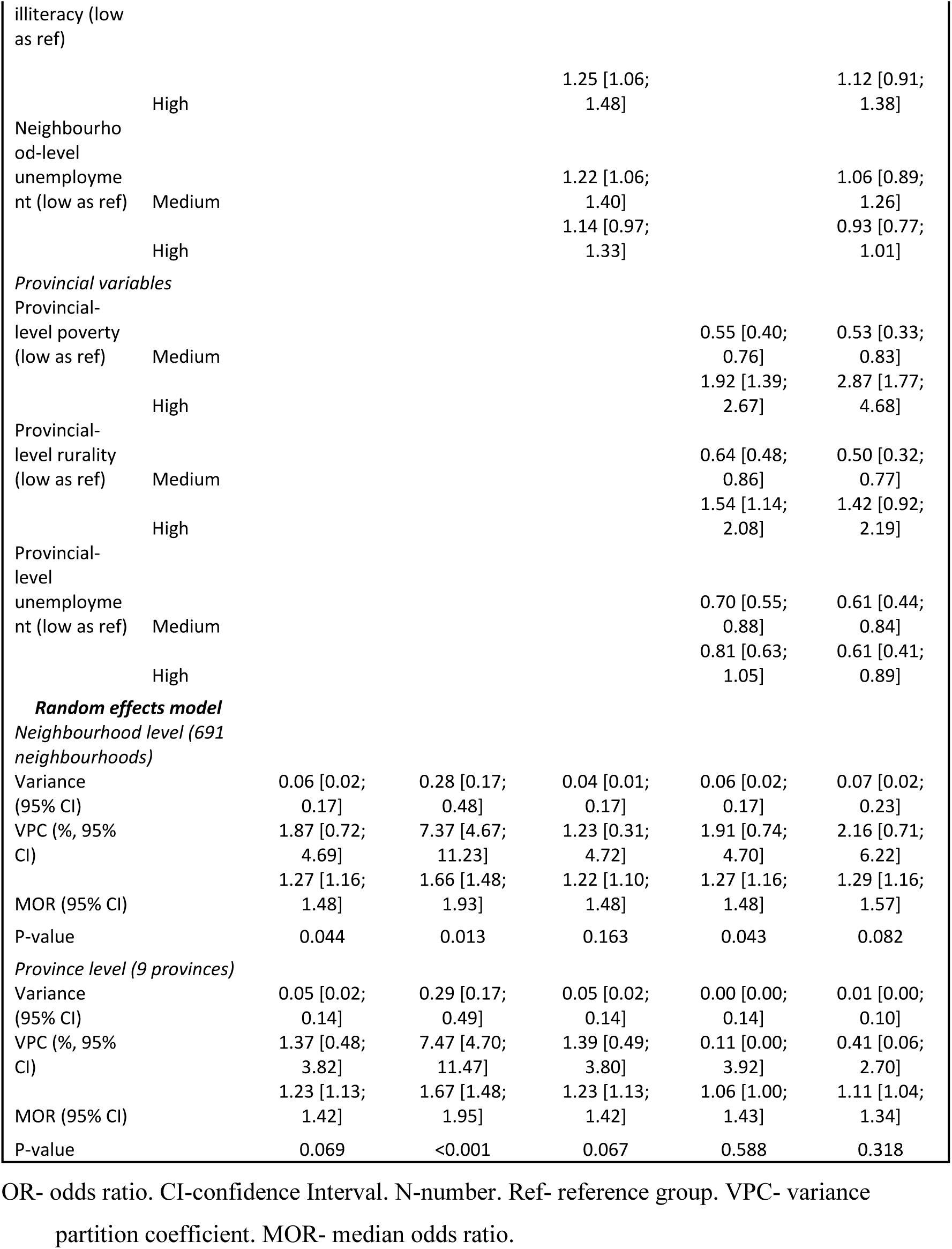
Individual, neighbourhood and province-level factors associated with multimorbidity identified by multilevel logistic regression models.

Men had 0.54 times the odds of being multimorbid compared to women (95%CI 0.45; 0.64). White and mixed-ancestry individuals had reduced odds of being multimorbid compared to black individuals (OR for white: 0.53, 95%CI 0.36; 0.79, OR for mixed-ancestry: 0.69 95%CI 0.51; 0.94). For every ten-year increase in an individual’s age, the odds of multimorbidity increased by 62% (OR 1.62, 95% CI 1.53; 1.71). Obese individuals had 45% increased odds of being multimorbid compared to individuals of a healthy weight (OR: 1.45, 95%CI 1.22; 1.72), and a medium level of sugary drink consumption was associated with reduced odds of multimorbidity compared to a low level of consumption (OR: 0.81, 95%CI 0.68; 0.96). Individuals exposed to smoke at work had 29% increased odds of multimorbidity (OR: 1.29, 95%CI 1.09; 1.54). In addition, having primary or secondary level education but not higher education, compared to no education, was associated with increased odds of multimorbidity (OR for primary: 1.32 95%CI 1.03;1.69, OR for secondary: 1.31 95%CI 1.01; 1.69). Lastly, individuals who were formerly married had 69% higher odds of multimorbidity, compared to those never married (OR: 1.69 95%CI 1.36; 2.10). No further associations were observed between individual level factors and multimorbidity.

Reduced odds of multimorbidity were observed for those in provinces with higher unemployment levels compared to provinces with the lowest unemployment levels (OR for middle unemployment: 0.61, 95%CI 0.44; 0.84, OR for least unemployment: 0.61, 95%CI 0.41; 0.89). In addition, individuals from a province with a medium level of rurality had reduced odds of multimorbidity compared to individuals from a province with a low level of rurality (OR: 0.50, 95%CI 0.32; 0.77). Individuals from provinces with a high level of poverty had increased odds of multimorbidity, whilst people from provinces with middle levels of poverty had reduced odds, compared to those from provinces with low levels of poverty (OR for richest: 2.87, 95%CI 1.77; 4.68, OR for middle: 0.53, 95%CI 0.33; 0.83). At the neighbourhood level, an association was observed between high levels of poverty and reduced odds of multimorbidity (OR: 0.77, 95%CI 0.62; 0.97).

### Random effects (measures of variation)

Modest variation in multimorbidity was observed across neighbourhoods and provinces in the empty model (σ^2^ for neighbourhoods: 0.06, 95%CI 0.02; 0.17, σ^2^ for provinces: 0.05, 95%CI 0.02; 0.14). In this model, 1.87% (VPC 95%CI 0.72; 4.69) and 1.37% (VPC 95%CI 0.48; 3.82) of the variation in odds of multimorbidity were consequent on neighbourhood and province level contextual factors respectively. In the final model containing all independent, neighbourhood and provincial covariates the odds of experiencing multimorbidity increased by 29% (MOR: 1.29, 95%CI 1.16; 1.57) on average for an individual moveing to a neighbourhood with a higher probability of multimorbidity. The VPCs for neighbourhood and province in model 5 were slightly increased and reduced, respectively, compared to model 1. There was no evidence of variation at the province level (P=0.32) and weak evidence of variation at the neighbourhood level (P=0.08).

### Sensitivity analysis

Of the 9,512 individuals (9,468 unweighted) from 734 neighbourhoods and 9 provinces, who completed the adult health module 33.6% were missing disease information on anaemia, 40.8% for HIV, 26.3% for hypertension and 38.9% for diabetes (see Supplementary file, Table S2). Therefore, there was considerable overlap between individuals missing information on different chronic conditions (43.8%, N=4,170). Individuals missing information on the outcome were more likely to be wealthy, highly educated, white and missing information on BMI than those with complete outcome data. Missingness was highest in Gauteng (53%) and lowest in North West (20.2%), see the Supplementary file, Table S3 and S4 for more information.

Results from sensitivity analysis using the larger sample of adults (9,512 weighted individuals), with multiple imputation to account for missing data can be found in Supplementary File, Figure S1, Table S5, S6, and S7. Almost a third (30.7%) of this population were found to be multimorbid, a similar pattern of chronic condition prevalence was seen as in the main analysis.

In the fully adjusted model, there was strong evidence of variation at the neighbourhood level (P<0.01) and weak evidence at the province level (P=0.07). Fixed effects associations were observed between increasing age, being educated to primary school level, being a woman, formerly married, obese, a medium level of new media access, exposure to smoke at work, and living in a province with a high level of poverty with multimorbidity, compared to their respective reference groups. Whilst individuals who worked skilled manual jobs, were white, from a province with a medium level of poverty and high level of unemployment had lower odds of multimorbidity, compared to their respective reference groups.

## Discussion

This research using SADHS 2016 data has estimated that almost half (44.6%) of adults in South Africa are multimorbid. The prevalence of multimorbidity ranged from 36.8% in Gauteng to 52.8% in Eastern Cape. After full adjustment, it was discovered that individuals who were older, women, black, obese, formerly married, consumed a medium quantity of sugary drinks, received education to primary or secondary school level, or exposed to smoke at work were all at an increased risk for multimorbidity. Individuals were less likely to have multimorbidity if they were from a province with a higher level of unemployment, medium level of rurality, or a neighbourhood with a high level of poverty. Contrastingly, those from a province with high levels of poverty were more likely to be multimorbid. Lastly, moderate geographical clustering of multimorbidity was observed, with a greater proportion of variance attributable to neighbourhood level factors than to province level factors.

### Prevalence in context

Estimates of the prevalence of multimorbidity in South Africa show substantial heterogeneity, ranging from 2.8% to 63.4% (Afshar et al., 2015; Alaba & Chola, 2013; Chidumwa et al., 2021; Garin et al., 2016; Roomaney et al., 2022; Weimann et al., 2016). These differences are likely the result of the inclusion of different disease conditions within multimorbidity, differing survey designs, analysis methods and age groups studied.

Analysis of the same dataset as used in this study (SADHS 2016) has previously found a prevalence of 20.7% (Roomaney et al., 2022). However, this study underestimated the true prevalence of multimorbidity due to being more restrictive with the conditions included (excluding chronic pain and cancer) and having misclassified individuals missing information on a disease (e.g. those who did not consent to biomarker measurement) as not having that disease. Sensitivity analyses using multiple imputation to account for missing data in this paper suggests a multimorbidity prevalence of 30.7%. This lower estimate is likely due to the individuals missing outcome data being more likely to be white, wealthy and highly educated, and therefore, less likely to be multimorbid.

Inclusion of different disease conditions has also led to considerably different prevalence estimates from the World Health Organisation’s study on Global AGEing and Adult Health (SAGE) in South Africa. Two sweeps from 2007/8 and 2014/15 reported a prevalence of 63.4% and 21.0% of multimorbidity in over 50s, respectively (Chidumwa et al., 2021; Garin et al., 2016). The latter study did not include 4 of the 11 conditions reported in the earlier study (obesity, cognitive impairment, edentulism and cataracts).

The lowest estimate of multimorbidity prevalence, from the South Africa National Income Dynamic Surveys 2012, utilised only four conditions of which three were self-reported (Weimann et al., 2016). Whilst, in certain groups such as the older, rural, and black population from the HAALSI study and in a survey of chronic patients attending health facilities in Tshwane prevalence of multimorbidity was especially high (70.9% and 98%, respectively) (Mkhwanazi et al., 2023; Wade et al., 2021).

### Fixed effect associations with multimorbidity

This paper provides further evidence that age, obesity, being formerly married and sex are associated with multimorbidity in South Africa (Alaba & Chola, 2013; Mkhwanazi et al., 2023; Weimann et al., 2016). However, surprisingly no associations were identified with health behaviours such as smoking, drinking and dietary health. This may be due to health behaviours altering once an individual is diagnosed with multiple chronic conditions, which this cross-sectional analysis could not capture (Chokshi et al., 2015). It is unclear why there was an association between medium but not high sugary drink consumption and multimorbidity; this was not identified in sensitivity analysis. In addition, exposure to smoke at work appears to be a novel association with multimorbidity.

The relationship between increasing education up to secondary level and reduced odds of multimorbidity could be explained by better access to and understanding of health information and healthcare resources which are important when preventing multimorbidity (Raghupathi & Raghupathi, 2020). Although it was notable that higher education was not linked to multimorbidity.

Neither new nor old media access, which were proxies for access to health information, were associated with multimorbidity. This may be due to individuals receiving more information from healthcare providers than through mass media.

Prior research has found increasing wealth to be linked to both an increased and decreased risk of multimorbidity in South Africa (Chang et al., 2019; Mkhwanazi et al., 2023; Weimann et al., 2016). Indeed, this research showed no clear pattern with individual wealth not being associated to multimorbidity, but increased neighbourhood poverty and medium province level poverty (compared to low province level poverty) linked to a reduction in multimorbidity, yet high province level poverty linked to an increase in multimorbidity. Sensitivity analysis results for neighbourhood and province variables were not consistent with these findings. More wealthy people may have greater access to health care, be more aware of their conditions, and therefore more likely to self-report issues. Nevertheless, they likely have a lower prevalence of multimorbidity risk factors (World Obesity Federation, 2020). Further investigation into these relationships is required, with a focus on the differing aspects that contribute to wealth in South Africa.

The link between residence (urban or rural) and multimorbidity in prior research is disparate, as were these findings (Alaba & Chola, 2013; Garin et al., 2016; Weimann et al., 2016). Lastly, a decreased risk of multimorbidity for individuals from provinces with higher levels of unemployment is interesting.

### Random effects in context

Moderate geographical clustering in multimorbidity was found in this study, including in the sensitivity analysis, with a greater proportion of the variance associated with neighbourhood (1.87% in the empty model and 2.16% in model 5) rather than province level factors (1.37% in the empty model and 0.41% in model 5). There was no evidence of clustering at the province level after taking into account observed variables. However, there was still weak evidence for residual variation at the neighbourhood level after full adjustment.

Prior research has noted the similarity in health states for individuals residing in the same geographical areas due to contextual peculiarities (Merlo et al., 2006). Similarities in multimorbidity within neighbourhoods may arise due to shared experiences, historical, political, geographical and cultural contexts and their proximity. However, shared factors in provinces with populations of at least 1.5 million may be limited (Maas & Hox, 2006). A previous study in Northern Ghana found 5.7% of the variance in multimorbidity to be explained by neighbourhood. The increased neighbourhood variance seen in Ghana could be explained by country differences or be due to that study not including regional level variation into their model.

### Policy implications

The health care model in South Africa is focused on managing single conditions, therefore individuals with multimorbidity are at risk of sub-optimal care for their conditions due to conflicting advice for discordant conditions and polypharmacy (Barnett et al., 2012; Mkhwanazi et al., 2023; Weimann et al., 2016). Policy and prevention measures need to be introduced to slow the growing population of multimorbid individuals in South Africa and to help manage current multimorbidity. This research suggests these prevention measures should be targeted at reducing exposure to smoke at work, increasing understanding and access to healthcare information through education and reducing unemployment, as well as considering neighbourhood level contexts.

### Strengths and limitations

This study reports nationally representative cross-sectional DHS data collected using a robust methodology and objective measurements of some diseases. Another strength is that only diseases believed to be current and chronic were included.

This paper was limited by the disease conditions asked about in the survey, meaning many diseases (e.g. mental health conditions) would have been missed. The prevalence of anaemia and diabetes may have been inflated due to a non-optimal testing approach (American Diabetes Association, 2021; National Department of Health et al., 2019). In addition, self-reporting of diseases may have led to under-ascertainment of the outcome and underestimating the prevalence of multimorbidity if individuals were unaware that they have a disease. This could be differential with respect to socioeconomic status, for example, if wealthy individuals were more likely to access health services, they would be more likely to self-report doctor diagnosed conditions, overestimating their risk of multimorbidity. These results should not be generalised to other countries where the contextual, area level and individual level factors may differ.

The results should not be interpreted causally. Firstly, this was a cross-sectional study forbidding the investigation of temporality, and limiting the ability to assess the length individuals were exposed to contextual factors. Furthermore, the fixed-effect coefficients from the models inherently adjusted for all other covariates, this may have led to adjustment for mediators or colliders and introduced bias to the results. Additionally, the province level results were based on a small sample size (N=9) so should be interpreted with caution. Also, a large number of individuals were excluded from this study due to missing information on the outcome which may have introduced selection bias. However, sensitivity analysis using multiple imputation was conducted to account for this missing data under the MAR approach. Lastly, there was likely to be residual confounding from unobserved (e.g. genetic, healthcare access and social capital) and poorly measured variables.

### Conclusions

This study estimates that 44.6% of adults in South Africa are prevalent for multimorbidity. Individuals who are older, women, formerly married, black, obese, consumed a medium amount of sugary drinks, received education to primary or secondary school level or exposed to smoke at work are at an increased risk of multimorbidity. Province level factors including poverty and unemployment, as well as neighbourhood level poverty were associated with multimorbidity. Some evidence of geographical clustering of multimorbidity was found, with greater variability due to neighbourhood level factors than province level factors. Therefore, policy and interventions that aim to tackle the burden of multimorbidity should consider the geographic clustering and shared drivers.

## Acknowledgements

The authors are grateful to Statistics South Africa (SSA), South African Medical Research Council (SAMRC), National Department of Health (NDoH) for providing the 2016 SADHS data for this analysis.

## Funding information

MH (Pre-doctoral Fellowship; NIHR302654) is funded by the National Institute for Health Research (NIHR). The views expressed in this publication are those of the author(s) and not necessarily those of the NIHR, NHS or the UK Department of Health and Social Care.

## Declarations of interest

The authors report there are no competing interests to declare.

## Data availability statement

SADHS data are available to download from: https://preview.dhsprogram.com/data/available-datasets.cfm.

## Notes

### Competing Interest Statement

The authors have declared no competing interest.

## References

Afshar, S., Roderick, P. J., Kowal, P., Dimitrov, B. D., & Hill, A. G. (2015). Multimorbidity and the inequalities of global ageing: A cross-sectional study of 28 countries using the World Health Surveys. BMC Public Health, 15(1), 1–10. 10.1186/S12889-015-2008-7/TABLES/4

Alaba, O., & Chola, L. (2013). The social determinants of multimorbidity in South Africa. International Journal for Equity in Health, 12(1), 1–10. 10.1186/1475-9276-12-63/TABLES/2

American Diabetes Association. (2021). 2. Classification and Diagnosis of Diabetes: Standards of Medical Care in Diabetes—2021. In Diabetes Care (Vol. 44, Issue Supplement_1). American Diabetes Association. 10.2337/DC21-S002

Ataguba, J. E., Akazili, J., & McIntyre, D. (2011). Socioeconomic-related health inequality in South Africa: evidence from General Household Surveys. International Journal for Equity in Health, 10. 10.1186/1475-9276-10-48

Barnett, K., Mercer, S. W., Norbury, M., Watt, G., Wyke, S., & Guthrie, B. (2012). Epidemiology of multimorbidity and implications for health care, research, and medical education: a cross-sectional study. Lancet (London, England), 380(9836), 37–43. 10.1016/S0140-6736(12)60240-2

Benebo, F. O., Schumann, B., & Vaezghasemi, M. (2018). Intimate partner violence against women in Nigeria: A multilevel study investigating the effect of women’s status and community norms. BMC Women’s Health, 18(1), 1–17. 10.1186/S12905-018-0628-7/FIGURES/3

Bolarinwa, O. A., Ahinkorah, B. O., Okyere, J., Seidu, A. A., & Olagunju, O. S. (2022). A multilevel analysis of prevalence and factors associated with female child marriage in Nigeria using the 2018 Nigeria Demographic and Health Survey data. BMC Women’s Health, 22(1), 1–11. 10.1186/S12905-022-01733-X/TABLES/2

Bradshaw, D., Groenewald, P., Laubscher, R., Nannan, N., Nojilana, B., Norman, R., Pieterse, D., Schneider, M., Bourne, D. E., Timæus, I. M., Dorrington, R., & Johnson, L. (2003). Initial burden of disease estimates for South Africa, 2000. South African Medical Journal, 93(9), 682–688. https://pubmed.ncbi.nlm.nih.gov/14635557/

Chang, A. Y., Gómez-Olivé, F. X., Payne, C., Rohr, J. K., Manne-Goehler, J., Wade, A. N., Wagner, R. G., Montana, L., Tollman, S., & Salomon, J. A. (2019). Chronic multimorbidity among older adults in rural South Africa. BMJ Global Health, 4(4), 1386. 10.1136/BMJGH-2018-001386

Chidumwa, G., Maposa, I., Corso, B., Minicuci, N., Kowal, P., Micklesfield, L. K., & Ware, L. J. (2021). Identifying co-occurrence and clustering of chronic diseases using latent class analysis: cross-sectional findings from SAGE South Africa Wave 2. BMJ Open, 11(1), e041604. 10.1136/BMJOPEN-2020-041604

Chokshi, D. A., El-Sayed, A. M., & Stine, N. W. (2015). J-Shaped Curves and Public Health. JAMA, 314(13), 1339–1340. 10.1001/JAMA.2015.9566

Chopra, M., & Sanders, D. (2004). From Apartheid to Globalisation: Health and Social Change in South Africa. Hygiea Internationalis : An Interdisciplinary Journal for the History of Public Health, 4(1), 153–174. 10.3384/hygiea.1403-8668.0441153

Chowdhury, S. R., Chandra Das, D., Sunna, T. C., Beyene, J., & Hossain, A. (2023). Global and regional prevalence of multimorbidity in the adult population in community settings: a systematic review and meta-analysis. EClinicalMedicine, 57. 10.1016/J.ECLINM.2023.101860

Consolazio, D., Murtas, R., Tunesi, S., Gervasi, F., Benassi, D., & Russo, A. G. (2021). Assessing the Impact of Individual Characteristics and Neighborhood Socioeconomic Status During the COVID-19 Pandemic in the Provinces of Milan and Lodi. International Journal of Health Services, 51(3), 311–324. 10.1177/0020731421994842/ASSET/IMAGES/LARGE/10.1177_0020731421994842-FIG1.JPEG

Corsi, D. J., Neuman, M., Finlay, J. E., & Subramanian, S. V. (2012). Demographic and health surveys: a profile. International Journal of Epidemiology, 41(6), 1602– 1613. 10.1093/IJE/DYS184

Due, P., Merlo, J., Harel-Fisch, Y., Damsgaard, M. T., Holstein, B. E., Hetland, J., Currie, C., Gabhainn, S. N., De Matos, M. G., & Lynch, J. (2009). Socioeconomic inequality in exposure to bullying during adolescence: A comparative, cross-sectional, multilevel study in 35 countries. American Journal of Public Health, 99(5), 907–914. 10.2105/AJPH.2008.139303

Fontes Marx, M., London, L., Harker, N., & Ataguba, J. E. (2021). Assessing Intertemporal Socioeconomic Inequalities in Alcohol Consumption in South Africa. Frontiers in Public Health, 9, 606050. 10.3389/FPUBH.2021.606050/BIBTEX

Frølich, A., Ghith, N., Schiøtz, M., Jacobsen, R., & Stockmarr, A. (2019). Multimorbidity, healthcare utilization and socioeconomic status: A register-based study in Denmark. PloS One, 14(8). 10.1371/JOURNAL.PONE.0214183

Garin, N., Koyanagi, A., Chatterji, S., Tyrovolas, S., Olaya, B., Leonardi, M., Lara, E., Koskinen, S., Tobiasz-Adamczyk, B., Ayuso-Mateos, J. L., & Haro, J. M. (2016). Global Multimorbidity Patterns: A Cross-Sectional, Population-Based, Multi-Country Study. The Journals of Gerontology: Series A, 71(2), 205–214. 10.1093/GERONA/GLV128

Ijaiya, M. A., Anjorin, S., & Uthman, O. A. (2022). Individual and contextual factors associated with childhood malnutrition: a multilevel analysis of the double burden of childhood malnutrition in 27 countries. Global Health Research and Policy, 7(1), 44. 10.1186/S41256-022-00276-W

Johnston, M. C., Crilly, M., Black, C., Prescott, G. J., & Mercer, S. W. (2019). Defining and measuring multimorbidity: a systematic review of systematic reviews. European Journal of Public Health, 29(1), 182–189. 10.1093/EURPUB/CKY098

Kuuire, V., Atuoye, K., Bisung, E., & Braimah, J. A. (2023). A Multilevel Analysis of Neighborhood Inequalities and Non-communicable Disease Multimorbidity in Ghana. 13–34. 10.1007/978-3-031-37565-1_2

Larsen, K., & Merlo, J. (2005). Appropriate Assessment of Neighborhood Effects on Individual Health: Integrating Random and Fixed Effects in Multilevel Logistic Regression. American Journal of Epidemiology, 161(1), 81–88. 10.1093/AJE/KWI017

Little, R. J. A., & Rubin, D. B. (2019). Statistical Analysis with Missing Data, 3rd Edition *| Wiley*. 464. https://www.wiley.com/en-us/Statistical+Analysis+with+Missing+Data%2C+3rd+Edition-p-9780470526798

Liyew, A. M., & Teshale, A. B. (2020). Individual and community level factors associated with anemia among lactating mothers in Ethiopia using data from Ethiopian demographic and health survey, 2016; A multilevel analysis. BMC Public Health, 20(1), 1–11. 10.1186/S12889-020-08934-9/TABLES/3

Loprinzi, P. D., Addoh, O., & Joyner, C. (2016). Multimorbidity, mortality, and physical activity. 10.1177/1742395316644306, 12(4), 272–280. 10.1177/1742395316644306

Maas, C. J. M., & Hox, J. J. (2006). Sufficient Sample Sizes for Multilevel Modeling. 10.1027/1614-2241.1.3.86, 1(3), 86–92. 10.1027/1614-2241.1.3.86

Makovski, T. T., Schmitz, S., Zeegers, M. P., Stranges, S., & van den Akker, M. (2019). Multimorbidity and quality of life: Systematic literature review and meta-analysis. Ageing Research Reviews, 53. 10.1016/J.ARR.2019.04.005

McGranahan, G., & Martine, G. (2012). Urbanisation and development: Policy lessons from the Brics’ experience. In Urban Growth in Emerging Economies: Lessons from the BRICS (pp. 1–14). 10.4324/9781315867878

Merlo, J., Chaix, B., Ohlsson, H., Beckman, A., Johnell, K., Hjerpe, P., Råstam, L., & Larsen, K. (2006). A brief conceptual tutorial of multilevel analysis in social epidemiology: using measures of clustering in multilevel logistic regression to investigate contextual phenomena. Journal of Epidemiology & Community Health, 60(4), 290–297. 10.1136/JECH.2004.029454

Merlo, J., Chaix, B., Yang, M., Lynch, J., & Råstam, L. (2005a). A brief conceptual tutorial of multilevel analysis in social epidemiology: linking the statistical concept of clustering to the idea of contextual phenomenon. Journal of Epidemiology and Community Health, 59(6), 443. 10.1136/JECH.2004.023473

Merlo, J., Chaix, B., Yang, M., Lynch, J., & Råstam, L. (2005b). A brief conceptual tutorial on multilevel analysis in social epidemiology: interpreting neighbourhood differences and the effect of neighbourhood characteristics on individual health. Journal of Epidemiology and Community Health, 59(12), 1022. 10.1136/JECH.2004.028035

Merlo, J., Yang, M., Chaix, B., Lynch, J., & Råstam, L. (2005). A brief conceptual tutorial on multilevel analysis in social epidemiology: investigating contextual phenomena in different groups of people. Journal of Epidemiology and Community Health, 59(9), 729. 10.1136/JECH.2004.023929

Mkhwanazi, T. W., Modjadji, P., Mokgalaboni, K., Madiba, S., & Roomaney, R. A. (2023). Multimorbidity, Treatment, and Determinants among Chronic Patients Attending Primary Health Facilities in Tshwane, South Africa. Diseases, 11(4). 10.3390/DISEASES11040129

National Department of Health, Statistics South Africa, South African Medical Research Council, & ICF. (2019). South Africa Demographic and Health Survey 2016.

Nunes, B. P., Flores, T. R., Mielke, G. I., Thumé, E., & Facchini, L. A. (2016). Multimorbidity and mortality in older adults: A systematic review and meta-analysis. Archives of Gerontology and Geriatrics, 67, 130–138. 10.1016/J.ARCHGER.2016.07.008

Oni, T., Youngblood, E., Boulle, A., Mcgrath, N., Wilkinson, R. J., & Levitt, N. S. (2015). Patterns of HIV, TB, and non-communicable disease multi-morbidity in peri-urban South Africa-a cross sectional study. 10.1186/s12879-015-0750-1

Raghupathi, V., & Raghupathi, W. (2020). The influence of education on health: An empirical assessment of OECD countries for the period 1995-2015. Archives of Public Health, 78(1), 1–18. 10.1186/S13690-020-00402-5/FIGURES/17

Roomaney, R. A., van Wyk, B., Cois, A., & Wyk, V. P. Van. (2022). One in five South Africans are multimorbid: An analysis of the 2016 demographic and health survey. PLOS ONE, 17(5), e0269081. 10.1371/JOURNAL.PONE.0269081

Roomaney, R. A., van Wyk, B., Turawa, E. B., & Pillay-van Wyk, V. (2021). Multimorbidity in South Africa: a systematic review of prevalence studies. BMJ Open, 11(10), e048676. 10.1136/BMJOPEN-2021-048676

Rubin, D. (2004). Multiple imputation for nonresponse in surveys. https://books.google.co.uk/books?hl=en&lr=&id=bQBtw6rx_mUC&oi=fnd&pg=PR24&ots=8PoJdP-XeN&sig=l35sET7zBnhtKf9-sLUTfAwNz5U

StataCorp. (2023). Stata Statistical Software: Release 18.

Sum, G., Salisbury, C., Koh, G. C. H., Atun, R., Oldenburg, B., Mcpake, B., Vellakkal, S., & Lee, J. T. (2019). Implications of multimorbidity patterns on health care utilisation and quality of life in middle-income countries: cross-sectional analysis. Journal of Global Health, 9(2). 10.7189/JOGH.09.020413

United States Agency for International Development. (2023). Guide to DHS Statistics. The Demographic and Health Surveys Program, 8. https://www.dhsprogram.com/Data/Guide-to-DHS-Statistics/index.cfm

Wade, A. N., Payne, C. F., Berkman, L., Chang, A., Gómez-Olivé, F. X., Kabudula, C., Kahn, K., Salomon, J. A., Tollman, S., Witham, M., & Davies, J. (2021). Multimorbidity and mortality in an older, rural black South African population cohort with high prevalence of HIV findings from the HAALSI Study. BMJ Open, 11(9), e047777. 10.1136/BMJOPEN-2020-047777

Weimann, A., Dai, D., & Oni, T. (2016). A cross-sectional and spatial analysis of the prevalence of multimorbidity and its association with socioeconomic disadvantage in South Africa: A comparison between 2008 and 2012. Social Science & Medicine, 163, 144–156. 10.1016/J.SOCSCIMED.2016.06.055

White, I. R., & Carlin, J. B. (2010). Bias and efficiency of multiple imputation compared with complete-case analysis for missing covariate values. Statistics in Medicine, 29(28), 2920–2931. 10.1002/SIM.3944

World Bank. (2024). South Africa *| Data*. https://data.worldbank.org/country/south-africa

World Health Organisation. (2023, March 16). Hypertension - Fact Sheet. https://www.who.int/news-room/fact-sheets/detail/hypertension

World Obesity Federation. (2020). Obesity: missing the 2025 global targets. In World Obesity Federation (Issue March). www.worldobesity.org

Xu, X., Mishra, G. D., & Jones, M. (2017). Mapping the global research landscape and knowledge gaps on multimorbidity: a bibliometric study. Journal of Global Health, 7(1). 10.7189/JOGH.07.010414

