## Supplementary files for "A multilevel analysis of the prevalence and factors associated with multimorbidity in South Africa using 2016 Demographic and Health Survey data"

*Table S1. Information on explanatory variables and their derivation*

| Variable | Information |
|---|---|
| Wealth index | Wealth index was characterised by DHS using principal component analysis of ownership of various household goods and other characteristics (i.e., type of flooring; source of water; availability of electricity; possession of durable consumer goods), split into quintiles (National Department of Health et al., 2019). For more information on this and other variables see: https://www.dhsprogram.com/publications/publication-fr337-dhs-final-reports.cfm?cssearch=16620_1 |
| Education level | Educational level was categorised into the highest educational level achieved, split into no education, primary-level education, secondary-level education, and higher-level education. Derived from ‘Highest education level’ variable. |
| Occupational status | Occupational status was split into eight categories: unemployed, professional/technical/management, clerical, agricultural, domestic services, sales and services, skilled manual, and unskilled manual. From the initial DHS categorisation ‘agriculture-self-employed’ and ‘agriculture – unskilled’ were combined, as were ‘household and domestic’ and ‘services’ to ensure sufficient data within categories. Derived from ‘Respondent currently working’ and ‘Respondent occupation (grouped)’ variables. |
| Health insurance | Whether individuals were covered by health insurance, yes/no (Y/N). Those missing information were coded as negative. Derived from ‘Covered by health insurance’ variable. |
| Marital status | Whether individuals were never in a union (married or living with partner), are currently in a union, or formerly in a union (widowed/ divorced/ separated). Derived from ‘Current marital status’ variable. |
| Age category | Age was categorised into ten-year bands from 15 to 64, and all those aged above 65 were banded together. Derived from ‘Current age’ variable. |
| Sex | Whether individuals were male or female. Derived from ‘Sex’ variable. |
| Ethnicity | Ethnicity categorised into black/African, white, mixed ancestry, and Asian/other. Derived from ‘Ethnicity’ variable. |
| Body mass index (BMI) | A digital scale and stadiometer were used to measure height and weight. According to WHO guidelines, BMI was categorised as follows: underweight as <18.5kg/m2, healthy weight between 18.5 and 24.9 kg/m2, overweight as between 25.0 and 29.9 kg/m2, and obesity as >30 kg/m2 (World Health Organisation, 2020). Women pregnant or who had given birth within two months were coded as missing, as BMI was not possible to ascertain. Derived from ‘Body Mass Index’, ‘Last birth to interview (months)’ and ‘Currently pregnant’ variables. |

| | |
|---|---|
| Dietary health | Six questions on dietary health were divided into four categories, combined together and split into low, medium and high categories. Derived from 'frequency eat fried foods', 'frequency eat fast foods', 'frequency eat packed chips', frequency east processed meat', 'types of fruit eaten yesterday', and 'types of vegetables eaten yesterday' variables. |
| Sugary drink intake | Five questions on sugary drink intake were combined and split into three categories (low, medium, high); 'Number of sugary drinks yesterday', 'Low amount of sugary drinks yesterday', and 'High amount of sugary drinks yesterday'. Derived from 'sugar-sweetened drinks yesterday', 'number of sugar-sweetened drinks of 200ml glass', 'number of sugar sweetened drinks of 330ml can or bottle', 'number of sugar-sweetened drinks of 500ml bottle', 'number of sugar-sweetened drinks of 1L bottle' and 'number of sugar-sweetened drinks of 2L bottle' variables. |
| Smoking status | 13 variables combined to determine whether individuals were current smokers (smoke manufactured cigarettes, hand-rolled cigarettes, pipe of tobacco, cigars and more tobacco-based products daily, weekly or less than weekly), ex-smokers (if smoked in the past but not currently) and never smokers. Derived from 'frequency currently smokes tobacco', 'frequency in the past smoked tobacco', 'on average respondent smokes daily: manufactured cigarettes', 'on average respondent smokes daily: hand roll cigarettes', 'on average respondent smokes daily: pipes full of tobacco', 'on average respondent smokes daily: cigars, cheroots, cigarillos', 'on average respondent smokes daily: water pipe sessions', 'on average respondent smokes daily: others', 'on average respondent smokes weekly: manufactured cigarettes', 'on average respondent smokes weekly: hand roll cigarettes', 'on average respondent smokes weekly: pipes full of tobacco', 'on average respondent smokes weekly: cigars, cheroots, cigarillos' and 'on average respondent smokes weekly: water pipe sessions' variables. |
| Alcohol drinking | Two questions were combined to determine whether individuals had drunk alcohol in the past 12 months (Y/N). Derived from 'ever consumed alcohol' and 'consumed alcohol in last 12 months' variables. |
| Exposure to smoke at work | Whether individuals were exposed to smoke at work (Y/N). Derived from 'ever worked in a place exposed to smoke' variable. |
| Access to old media | Responses to three questions on frequency of watching television, listening to radio, and reading newspaper or magazine, were combined and split into low, medium and high categories. Derived from 'frequency of reading newspaper or magazine', 'frequency of listening to radio' and 'frequency of watching television' variables. |
| Access to new media | Responses to two questions on whether owns a mobile phone and frequency of internet use in the preceding month were combined and split into low, medium and high categories. Derived from 'frequency of using internet last month' and 'owns a mobile phone' variables. |
| Neighbourhood-level poverty | Neighbourhoods were defined as respondents from clusters of households which serve as the PSU within the DHS. Defined as the proportion of individuals living in poverty for each neighbourhood. This was split into three categories (low, medium and high) with low as the reference group and calculated with the larger adult-health sample (N= 9,512) to include more contextual information. Derived from 'Wealth index' variable. |

| | |
|---|---|
| Neighbourhood-level rurality | Neighbourhoods were defined as respondents from clusters of households which serve as the PSU within the DHS. Defined as whether a neighbourhood resided in an urban or rural area and calculated with the larger adult-health sample (N= 9,512) with urban as the reference group to include more contextual information. Derived from 'place of residence' variable. |
| Neighbourhood-level illiteracy | Neighbourhoods were defined as respondents from clusters of households which serve as the PSU within the DHS. Defined as the proportion of individuals illiterate in each neighbourhood. This was split into three categories (low, medium and high) with low as the reference group and calculated with the larger adult-health sample (N= 9,512) to include more contextual information. Derived from 'literacy' variable. |
| Neighbourhood-level unemployment | Neighbourhoods were defined as respondents from clusters of households which serve as the PSU within the DHS. Defined as the proportion of individuals unemployed in each neighbourhood. This was split into three categories (low, medium and high) with low as the reference group and calculated with the larger adult-health sample (N= 9,512) to include more contextual information. Derived from 'Respondent currently working' and 'Respondent occupation (grouped)' variables. |
| Provincial-level poverty | Defined as the proportion of individuals living in poverty for each province. This was split into three categories (low, medium and high) with low as the reference group and calculated with the larger adult-health sample (N= 9,512) to include more contextual information. Derived from 'Wealth index' variable. |
| Provincial-level rurality (low as ref) | Defined as the proportion of individuals living in rural areas for each province. This was split into three categories (low, medium and high) with low as the reference group and calculated with the larger adult-health sample (N= 9,512) to include more contextual information. Derived from 'place of residence' variable. |
| Provincial-level unemployment (low as ref) | Defined as the proportion of individuals unemployed for each province. This was split into three categories (low, medium and high) with low as the reference group and calculated with the larger adult-health sample (N= 9,512) to include more contextual information. Derived from 'Respondent currently working' and 'Respondent occupation (grouped)' variables. |

*Table S2. Missingness of chronic diseases for adults that completed the adult health sample in SADHS 2016 (N=9,512)*

| **Chronic disease** | **Missing** | |
|---|---|---|
| | **N** | **%** |
| HIV | 3885 | 40.8 |
| Diabetes | 3704 | 38.9 |
| Anaemia | 3193 | 33.6 |
| Hypertension | 2502 | 26.3 |
| Cancer | 0 | 0.0 |
| Stroke | 0 | 0.0 |
| Tuberculosis | 0 | 0.0 |
| Chronic bronchitis | 0 | 0.0 |
| Asthma | 0 | 0.0 |
| Heart attack | 0 | 0.0 |
| High blood cholesterol | 0 | 0.0 |
| Chronic pain | 0 | 0.0 |

Adjusted for sample weight, stratification and clustering.

*Table S3. Summary of the SADHS 2016 sample by multimorbidity status including those missing data on outcome*

| | | Overall | People without multimorbidity | People with multimorbidity | People missing multimorbidity |
|---|---|---|---|---|---|
| Total, N (% col) | | 9,512 (100.0%) | 2,960 (31.1%) | 2,382 (25.0%) | 4,171 (43.8%) |
| *Individual socio-economic variables* | | | | | |
| Wealth index, N (% col) | Poorest | 1,748 (18.4%) | 628 (21.2%) | 500 (21.0%) | 621 (14.9%) |
| | Poorer | 1,827 (19.2%) | 588 (19.9%) | 475 (19.9%) | 764 (18.3%) |
| | Middle | 1,963 (20.6%) | 628 (21.2%) | 521 (21.9%) | 813 (19.5%) |
| | Richer | 1,917 (20.2%) | 569 (19.2%) | 470 (19.7%) | 878 (21.1%) |
| | Richest | 2,057 (21.6%) | 546 (18.4%) | 416 (17.5%) | 1,095 (26.3%) |
| Education, N (% col) | No education | 711 (7.5%) | 154 (5.2%) | 294 (12.3%) | 263 (6.3%) |
| | Primary | 1,535 (16.1%) | 437 (14.8%) | 584 (24.5%) | 514 (12.3%) |
| | Secondary | 6,145 (64.6%) | 2,018 (68.2%) | 1,324 (55.6%) | 2,803 (67.2%) |
| | Higher | 1,122 (11.8%) | 351 (11.8%) | 180 (7.6%) | 591 (14.2%) |
| Occupation, N (% col) | Unemployed | 5,853 (61.5%) | 1,856 (62.7%) | 1,609 (67.5%) | 2,389 (57.3%) |
| | Professional/technical | 674 (7.1%) | 195 (6.6%) | 108 (4.6%) | 370 (8.9%) |
| | Clerical | 356 (3.7%) | 102 (3.4%) | 65 (2.7%) | 189 (4.5%) |
| | Agricultural | 122 (1.3%) | 40 (1.3%) | 29 (1.2%) | 53 (1.3%) |
| | Domestic service | 191 (2.0%) | 46 (1.6%) | 54 (2.3%) | 91 (2.2%) |
| | Sales and services | 521 (5.5%) | 161 (5.4%) | 113 (4.7%) | 248 (5.9%) |
| | Skilled manual | 670 (7.0%) | 246 (8.3%) | 114 (4.8%) | 310 (7.4%) |
| | Unskilled manual | 689 (7.2%) | 177 (6.0%) | 222 (9.3%) | 290 (7.0%) |
| | Missing | 436 (4.6%) | 137 (4.6%) | 68 (2.8%) | 231 (5.5%) |
| Have health insurance, N (% col) | Yes | 1,094 (11.5%) | 321 (10.8%) | 165 (6.9%) | 608 (14.6%) |
| *Individual socio-demographic variables* | | | | | |
| Mean age, years (SD) | | 40.9 (17.2) | 35.5 (15.7) | 49.1 (17.0) | 40.0 (16.6) |
| Sex, N (% col) | Men | 3,821 (40.2%) | 1,321 (44.6%) | 652 (27.4%) | 1,848 (44.3%) |
| Ethnicity, N (% col) | Black | 7,962 (83.7%) | 2,543 (85.9%) | 2,069 (86.9%) | 3,350 (80.3%) |
| | White | 563 (5.9%) | 172 (5.8%) | 106 (4.5%) | 284 (6.8%) |

| | | | | | |
|---|---|---|---|---|---|
| | Mixed | 794 (8.3%) | 210 (7.1%) | 169 (7.1%) | 415 (10.0%) |
| | Other | 194 (2.0%) | 34 (1.2%) | 38 (1.6%) | 121 (2.9%) |
| Marital status, N (% col) | Never married | 4,478 (47.1%) | 1,658 (56.0%) | 845 (35.5%) | 1,975 (47.3%) |
| | Formerly married | 1,164 (12.2%) | 217 (7.3%) | 494 (20.7%) | 453 (10.9%) |
| | Currently married | 3,870 (40.7%) | 1,085 (36.7%) | 1,043 (43.8%) | 1,742 (41.8%) |
| Old media access, N (% col) | Low | 3,432 (36.1%) | 1,094 (37.0%) | 997 (41.8%) | 1,341 (32.2%) |
| | Medium | 3,440 (36.2%) | 1,090 (36.8%) | 841 (35.3%) | 1,509 (36.2%) |
| | High | 2,641 (27.8%) | 775 (26.2%) | 545 (22.9%) | 1,321 (31.7%) |
| New media access, N (% col) | Low | 920 (9.7%) | 263 (8.9%) | 282 (11.8%) | 375 (9.0%) |
| | Medium | 5,055 (53.1%) | 1,420 (48.0%) | 1,581 (66.4%) | 2,054 (49.3%) |
| | High | 3,537 (37.2%) | 1,277 (43.1%) | 519 (21.8%) | 1,741 (41.7%) |
| *Individual health variables* | | | | | |
| Body mass index, N (% col) | Underweight | 326 (3.4%) | 151 (5.1%) | 72 (3.0%) | 104 (2.5%) |
| | Healthy weight | 2,822 (29.7%) | 1,304 (44.0%) | 665 (27.9%) | 853 (20.5%) |
| | Overweight | 1,830 (19.2%) | 743 (25.1%) | 620 (26.0%) | 467 (11.2%) |
| | Obese | 2,215 (23.3%) | 668 (22.6%) | 974 (40.9%) | 573 (13.7%) |
| | Missing | 2,318 (24.4%) | 94 (3.2%) | 50 (2.1%) | 2,174 (52.1%) |
| Smoking status, N (% col) | Never smoker | 7,109 (74.7%) | 2,165 (73.1%) | 1,887 (79.2%) | 3,058 (73.3%) |
| | Current smoker | 1,983 (20.8%) | 659 (22.3%) | 384 (16.1%) | 940 (22.5%) |
| | Former smoker | 420 (4.4%) | 136 (4.6%) | 111 (4.7%) | 172 (4.1%) |
| Drink alcohol, N (% col) | Yes | 3,220 (33.8%) | 1,144 (38.6%) | 635 (26.7%) | 1,441 (34.5%) |
| Dietary health, N (% col) | High | 4,358 (45.8%) | 1,287 (43.5%) | 1,246 (52.3%) | 1,824 (43.7%) |
| | Medium | 2,964 (31.2%) | 924 (31.2%) | 724 (30.4%) | 1,316 (31.6%) |
| | Poor | 2,190 (23.0%) | 749 (25.3%) | 412 (17.3%) | 1,030 (24.7%) |
| Sugary drink intake, N (% col) | Low | 6,138 (64.5%) | 1,789 (60.4%) | 1,693 (71.1%) | 2,656 (63.7%) |
| | Medium | 1,822 (19.2%) | 612 (20.7%) | 356 (14.9%) | 854 (20.5%) |
| | High | 1,552 (16.3%) | 559 (18.9%) | 333 (14.0%) | 660 (15.8%) |
| Exposure to smoke at work, N (% col) | Yes | 1,924 (20.2%) | 574 (19.4%) | 508 (21.3%) | 843 (20.2%) |

N- number. % Col- column percentages. SD- standard deviation. Adjusted for sample weight, stratification and clustering.

Table S4. Summary of the SADHS 2016 sample by multimorbidity status and province including those missing data on outcome

| | | Overall | | People without multimorbidity | | People with multimorbidity | | People missing multimorbidity | |
|---|---|---|---|---|---|---|---|---|---|
| | | N | % col | N | % | N | % | N | % |
| Total | | 9,512 | 100.0 | 2,959 | 31.1 | 2,382 | 25.0 | 4,170 | 43.8 |
| Region | Western Cape | 1105 | 11.6 | 286 | 25.9 | 248 | 22.5 | 571 | 51.6 |
| | Eastern Cape | 1101 | 11.6 | 377 | 34.2 | 422 | 38.3 | 303 | 27.5 |
| | Northern Cape | 194 | 2.0 | 47 | 24.2 | 45 | 23.3 | 102 | 52.5 |
| | Free state | 485 | 5.1 | 192 | 39.5 | 164 | 33.8 | 129 | 26.7 |
| | KwaZulu-Natal | 1713 | 18.0 | 477 | 27.8 | 441 | 25.8 | 795 | 46.4 |
| | Northwest | 670 | 7.0 | 309 | 46.2 | 225 | 33.6 | 135 | 20.2 |
| | Gauteng | 2588 | 27.2 | 769 | 29.7 | 448 | 17.3 | 1371 | 53.0 |
| | Mpumalanga | 733 | 7.7 | 184 | 25.1 | 201 | 27.4 | 348 | 47.4 |
| | Limpopo | 922 | 9.7 | 318 | 34.5 | 187 | 20.3 | 417 | 45.2 |

N- number. % Col- column percentages. SD- standard deviation. Adjusted for sample weight, stratification and clustering.

*Table S5. Summary of the sample using multiple imputation by multimorbidity status*

| | | People without multimorbidity | | People with multimorbidity | |
|---|---|---|---|---|---|
| | | % | N | % | N |
| Total, N (% col) | | 69.3 | 6596 | 30.7 | 2916 |
| *Individual socio-economic variables* | | | | | |
| Wealth index, N (% col) | Poorest | 17.8 | 1238 | 19.7 | 575 |
| | Poorer | 19.0 | 1324 | 19.6 | 571 |
| | Middle | 20.4 | 1416 | 21.3 | 620 |
| | Richer | 20.1 | 1399 | 20.2 | 590 |
| | Richest | 22.7 | 1579 | 19.2 | 560 |
| Education, N (% col) | No education | 5.3 | 366 | 12.5 | 364 |
| | Primary | 12.8 | 887 | 23.8 | 693 |
| | Secondary | 68.5 | 4767 | 55.7 | 1624 |
| | Higher | 13.4 | 935 | 8.1 | 235 |
| Occupation, N (% col) | Unemployed | 61.7 | 4289 | 69.2 | 2019 |
| | Professional/technical | 8.5 | 591 | 5.3 | 156 |
| | Clerical | 4.4 | 309 | 2.8 | 83 |
| | Agricultural | 1.5 | 105 | 1.2 | 34 |
| | Domestic service | 1.9 | 130 | 2.5 | 74 |
| | Sales and services | 6.3 | 442 | 4.7 | 138 |
| | Skilled manual | 8.9 | 617 | 4.7 | 138 |
| | Unskilled manual | 6.8 | 473 | 9.4 | 275 |
| Have health insurance, N (% col) | Yes | 13.5 | 940 | 7.0 | 203 |
| *Individual socio-demographic variables* | | | | | |
| Mean age, years (SD) | | 37.1 (0.3) | | 49.6 (0.6) | |
| Sex, N (% col) | Men | 45.6 | 3171 | 27.9 | 814 |
| Ethnicity, N (% col) | Black | 83.4 | 5803 | 84.4 | 2460 |
| | White | 5.9 | 411 | 5.9 | 173 |
| | Mixed ancestry | 8.6 | 597 | 7.8 | 228 |

| | | | | | |
|---|---|---|---|---|---|
| | Other | 2.1 | 146 | 1.9 | 55 |
| Marital status, N (% col) | Never married | 52.1 | 3626 | 35.7 | 1040 |
| | Formerly married | 8.4 | 581 | 21.0 | 613 |
| | Currently married | 39.5 | 2750 | 43.3 | 1263 |
| Old media access, N (% col) | Low | 33.8 | 2352 | 41.2 | 1202 |
| | Medium | 36.2 | 2521 | 36.0 | 1049 |
| | High | 29.9 | 2083 | 22.8 | 666 |
| New media access, N (% col) | Low | 8.7 | 603 | 11.9 | 348 |
| | Medium | 47.7 | 3319 | 65.4 | 1907 |
| | High | 43.6 | 3033 | 22.7 | 661 |
| *Individual health variables* | | | | | |
| Body mass index, N (% col)* | Underweight | 5.3 | 369 | 3.1 | 91 |
| | Healthy weight | 43.3 | 3010 | 29.1 | 849 |
| | Overweight | 25.2 | 1752 | 25.9 | 757 |
| | Obese | 26.2 | 1825 | 41.8 | 1220 |
| Smoking status, N (% col) | Never smoker | 72.9 | 5071 | 78.9 | 2301 |
| | Current smoker | 23.0 | 1599 | 16.0 | 467 |
| | Former smoker | 4.1 | 287 | 5.1 | 148 |
| Drink alcohol, N (% col) | Yes | 37.0 | 2575 | 26.7 | 778 |
| Dietary health, N (% col) | High | 42.8 | 2974 | 52.7 | 1537 |
| | Medium | 31.7 | 2202 | 30.0 | 876 |
| | Poor | 25.6 | 1780 | 17.3 | 503 |
| Sugary drink intake, N (% col) | Low | 61.8 | 4298 | 70.7 | 2062 |
| | Medium | 20.9 | 1453 | 15.2 | 445 |
| | High | 17.3 | 1205 | 14.0 | 409 |
| Exposure to smoke at work, N (% col) | Yes | 19.3 | 1341 | 22.4 | 652 |

N- number. % Col- column percentages. SD- standard deviation. Adjusted for sample weight, stratification and clustering. *BMI missing 214 people

*Table S6. Description of the sample using multiple imputation by region*

| Province | Number of people N | (% col) | Age Mean | SD | Multimorbid N | % row | Poverty level, % row | Rurality level, % row | Illiteracy level, % row | Unemployment level, % row |
|---|---|---|---|---|---|---|---|---|---|---|
| Total | 9512 | 100.0 | 40.9 | 0.3 | 2916 | 100.0 | 20.0 | 37.5 | 10.1 | 64.6 |
| Western Cape | 1106 | 11.6 | 44.6 | 1.0 | 336 | 30.4 | 4.8 | 3.7 | 4.9 | 55.4 |
| Eastern Cape | 1101 | 11.6 | 43.1 | 0.7 | 475 | 43.1 | 38.9 | 55.3 | 14.1 | 70.7 |
| Northern Cape | 194 | 2.0 | 42.6 | 0.6 | 65 | 33.2 | 11.1 | 26.6 | 9.0 | 67.1 |
| Free state | 485 | 5.1 | 41.4 | 0.6 | 180 | 37.1 | 7.1 | 11.5 | 7.7 | 72.3 |
| KwaZulu-Natal | 1713 | 18.0 | 39.8 | 0.8 | 527 | 30.8 | 28.4 | 52.8 | 10.6 | 70.2 |
| North West | 670 | 7.0 | 40.2 | 1.3 | 239 | 35.7 | 14.6 | 55.7 | 11.8 | 59.4 |
| Gauteng | 2588 | 27.2 | 39.1 | 0.8 | 590 | 22.8 | 15.4 | 10.7 | 6.1 | 60.8 |
| Mpumalanga | 733 | 7.7 | 38.6 | 0.7 | 267 | 36.4 | 19.6 | 63.9 | 14.8 | 62.4 |
| Limpopo | 922 | 9.7 | 42.6 | 0.7 | 237 | 25.7 | 25.7 | 85.0 | 17.9 | 69.7 |

Adjusted for sample weight, stratification and clustering.

*Figure S1. Prevalence of individual chronic diseases in the sample using multiple imputation*

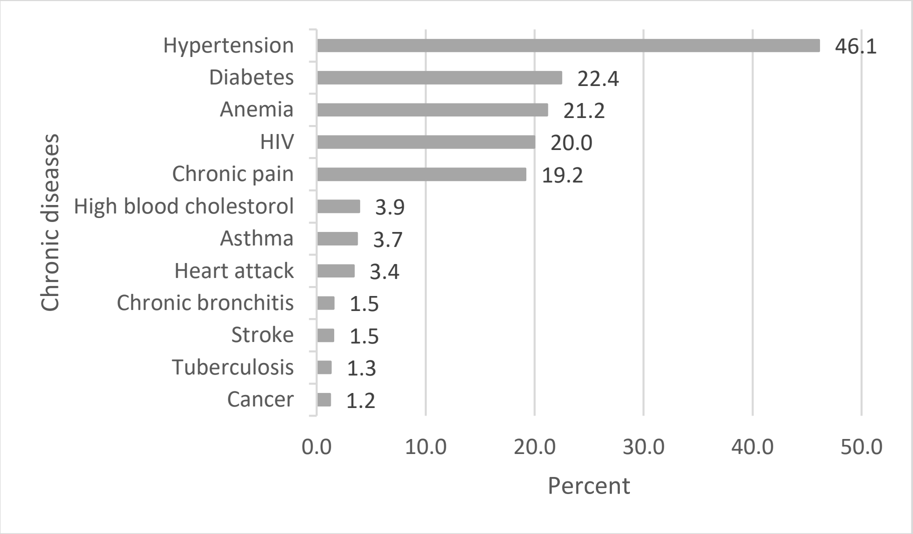

Adjusted for sample weight, stratification and clustering.

*Table S7. Individual, neighbourhood, and regional-level factors associated with multimorbidity identified by multilevel logistic regression models in the sample using multiple imputation*

| | | Model 1 OR (95% CI) (N=9,468) | Model 2 OR (95% CI) (N=9,254) | Model 3 OR (95% CI) (N=9,468) | Model 4 OR (95% CI) (N=9,468) | Model 5 OR (95% CI) (N=9,254) |
|---|---|---|---|---|---|---|
| ***Fixed effect model*** | | | | | | |
| *Individual socio-economic variables* | | | | | | |
| Wealth index (poorest as ref) | Poorer | | 1.01 [0.98; 1.04] | | | 1 [0.97; 1.03] |
| | Middle | | 1 [0.97; 1.03] | | | 1 [0.96; 1.03] |
| | Richer | | 0.99 [0.96; 1.03] | | | 0.99 [0.95; 1.03] |
| | Richest | | 0.96 [0.92; 1] | | | 0.96 [0.91; 1.01] |
| Education (none as ref) | Primary | | 1.05 [1.01; 1.08] | | | 1.05 [1.01; 1.08] |
| | Secondary | | 1.03 [0.99; 1.06] | | | 1.03 [0.99; 1.07] |
| | Higher | | 0.98 [0.93; 1.03] | | | 0.98 [0.93; 1.03] |
| Occupation (unemployed as ref) | Professional/ technical | | 0.97 [0.93; 1.01] | | | 0.97 [0.93; 1.01] |
| | Clerical | | 0.96 [0.91; 1.01] | | | 0.96 [0.91; 1.01] |
| | Agricultural | | 0.98 [0.92; 1.05] | | | 0.98 [0.92; 1.05] |
| | Domestic service | | 1.04 [0.97; 1.11] | | | 1.04 [0.97; 1.11] |
| | Sales and services | | 0.98 [0.94; 1.02] | | | 0.98 [0.94; 1.02] |
| | Skilled manual | | 0.95 [0.91; 0.98] | | | 0.95 [0.91; 0.98] |
| | Unskilled manual | | 1.01 [0.98; 1.05] | | | 1.01 [0.98; 1.05] |
| Have health insurance (no as ref) | Yes | | 0.98 [0.95; 1.02] | | | 0.98 [0.95; 1.02] |
| *Individual socio-demographic variables* | | | | | | |
| Age category (5-year groups) | | | 1.09 [1.08; 1.1] | | | 1.09 [1.08; 1.1] |
| Sex (women as ref) | Men | | 0.89 [0.87; 0.91] | | | 0.89 [0.87; 0.91] |
| Ethnicity (black as ref) | White | | 0.92 [0.87; 0.96] | | | 0.92 [0.87; 0.97] |
| | Mixed ancestry | | 0.95 [0.91; 0.99] | | | 0.94 [0.9; 0.98] |
| | Other | | 0.94 [0.86; 1.02] | | | 0.94 [0.87; 1.03] |

| | | | | |
|---|---|---|---|---|
| Marital status (never married as ref) | Formerly married | 1.06 [1.03; 1.1] | | 1.06 [1.03; 1.1] |
| | Currently married | 1 [0.98; 1.02] | | 1 [0.98; 1.02] |
| Old media access (low as ref) | Medium | 1.01 [0.99; 1.03] | | 1.01 [0.99; 1.03] |
| | High | 0.99 [0.96; 1.01] | | 0.99 [0.96; 1.01] |
| New media access (low as ref) | Medium | 1.04 [1.01; 1.07] | | 1.04 [1.01; 1.07] |
| | High | 1.01 [0.98; 1.05] | | 1.01 [0.98; 1.05] |
| *Individual health variables* | | | | |
| Body mass index (healthy weight as ref) | Underweight | 1.02 [0.98; 1.07] | | 1.02 [0.98; 1.07] |
| | Overweight | 1.02 [0.99; 1.04] | | 1.02 [0.99; 1.04] |
| | Obese | 1.07 [1.04; 1.1] | | 1.07 [1.04; 1.09] |
| Smoking status (never smoker as ref) | Current smoker | 0.99 [0.96; 1.01] | | 0.99 [0.96; 1.01] |
| | Former smoker | 1.04 [0.99; 1.08] | | 1.04 [0.99; 1.08] |
| Drink alcohol (no as ref) | Yes | 1.01 [0.99; 1.03] | | 1.01 [0.99; 1.03] |
| Dietary health (high as ref) | Medium | 1 [0.98; 1.02] | | 1 [0.98; 1.02] |
| | Poor | 0.98 [0.96; 1.01] | | 0.99 [0.96; 1.01] |
| Sugary drink intake (low as ref) | Medium | 0.98 [0.96; 1.01] | | 0.98 [0.96; 1.01] |
| | High | 0.99 [0.97; 1.01] | | 0.99 [0.97; 1.02] |
| Exposure to smoke at work (no as ref) | Yes | 1.07 [1.05; 1.1] | | 1.07 [1.05; 1.1] |
| *Neighbourhood variables* | | | | |
| Neighbourhood-level poverty (low as ref) | Medium | | 1 [0.96; 1.03] | 1.01 [0.98; 1.04] |
| | High | | 0.97 [0.95; 1] | 0.98 [0.95; 1.01] |
| Neighbourhood-level rurality (urban as ref) | Rural | | 1.05 [1.02; 1.08] | 1.02 [0.99; 1.05] |

| | | | | | | |
|---|---|---|---|---|---|---|
| Neighbourhood-level illiteracy (low as ref) | Medium | | | 1.03 [1.01; 1.06] | | 1.02 [0.99; 1.05] |
| | High | | | 1.04 [1.01; 1.08] | | 1.02 [0.99; 1.06] |
| Neighbourhood-level unemployment (low as ref) | Medium | | | 1.05 [1.02; 1.08] | | 1.01 [0.99; 1.04] |
| | High | | | 1.05 [1.02; 1.08] | | 0.99 [0.96; 1.02] |
| *Provincial variables* | | | | | | |
| Provincial-level poverty (low as ref) | Medium | | | | 0.9 [0.86; 0.95] | 0.91 [0.86; 0.97] |
| | High | | | | 1.04 [0.98; 1.1] | 1.1 [1.03; 1.18] |
| Provincial-level rurality (low as ref) | Medium | | | | 1.01 [0.96; 1.06] | 0.95 [0.9; 1.01] |
| | High | | | | 1.1 [1.05; 1.16] | 1.05 [0.99; 1.11] |
| Provincial-level unemployment (low as ref) | Medium | | | | 0.95 [0.91; 0.98] | 0.92 [0.88; 0.96] |
| | High | | | | 1.04 [1; 1.09] | 0.98 [0.94; 1.04] |
| ***Random effects model*** | | | | | | |
| *Neighbourhood level (734 neighbourhoods)* | | | | | | |
| Variance (95% CI) | | 0.07 [0.06; 0.08] | 0.07 [0.06; 0.09] | 0.07 [0.06; 0.09] | 0.08 [0.07; 0.10] | 0.07 [0.06; 0.09] |
| VPC (%, 95% CI) | | 2.41 [2.03; 2.85] | 2.20 [1.83; 2.61] | 2.09 [1.69; 2.58] | 2.46 [2.07; 2.80] | 2.17 [1.80; 2.60] |
| MOR (95% CI) | | 1.30 [1.27; 1.33] | 1.30 [1.27; 1.33] | 1.24 [1.19; 1.33] | 1.32 [1.29; 1.35] | 1.29 [1.26; 1.33] |
| P-value | | <0.001 | <0.001 | <0.001 | <0.001 | <0.001 |
| *Province level (9 provinces)* | | | | | | |
| Variance (95% CI) | | 0.05 [0.03; 0.08] | 0.05 [0.03; 0.08] | 0.05 [0.03; 0.09] | 0.01 [0.00; 0.17] | 0.01 [0.00; 0.05] |
| VPC (%, 95% CI) | | 1.57 [0.95; 2.56] | 1.37 [0.83; 2.26] | 1.54 [0.94; 2.51] | 0.23 [0.10; 4.88] | 0.38 [0.11; 1.37] |
| MOR (95% CI) | | 1.25 [1.17; 1.31] | 1.23 [1.17; 1.31] | 1.29 [1.26; 1.33] | 1.09 [1.02; 1.49] | 1.11 [1.06; 1.23] |
| P-value | | <0.001 | <0.001 | <0.001 | 0.53 | 0.07 |

OR- odds ratio. CI-confidence Interval. Ref- reference group. VPC – variance partition coefficient. MOR- median odds ratio.
